# The differential impact of physical distancing strategies on social contacts relevant for the spread of COVID-19: Evidence from a multi-country survey

**DOI:** 10.1101/2020.05.15.20102657

**Authors:** Emanuele Del Fava, Jorge Cimentada, Daniela Perrotta, André Grow, Francesco Rampazzo, Sofia Gil-Clavel, Emilio Zagheni

## Abstract

Physical distancing measures are intended to mitigate the spread of COVID-19, even though their impact on social contacts and disease transmission remains unclear. Obtaining timely data on social contact patterns can help to assess the impact of such protective measures. We conducted an online opt-in survey based on targeted Facebook advertising campaigns across seven European countries (Belgium, France, Germany, Italy, Netherlands, Spain, United Kingdom (UK)) and the United States (US), achieving a sample of 53,708 questionnaires in the period March 13–April 13, 2020. Post-stratification weights were produced to correct for biases. Data on social contact numbers, as well as on protective behaviour and perceived level of threat were collected and used to the expected net reproduction number by week, *R*_*t*_, with respect to pre-pandemic data. Compared to social contacts reported prior to COVID-19, in mid-April daily social contact numbers had decreased between 49% in Germany and 83% in Italy, ranging from below three contacts per day in France, Spain, and the UK up to four in Germany and the Netherlands. Such reductions were sufficient to bring *R*_*t*_ to one or even below in all countries, except Germany. Evidence from the US and the UK showed that the number of daily social contacts mainly decreased after governments issued the first physical distancing guidelines. Finally, although contact numbers decreased uniformly across age groups, older adults reported the lowest numbers of contacts, indicating higher levels of protection. We provided a comparable set of statistics on social contact patterns during the COVID-19 pandemic for eight high-income countries, disaggregated by week. As these estimates offer a more grounded alternative to the theoretical assumptions often used in epidemiological models, the scientific community could draw on this information for developing more realistic epidemic models of COVID-19.

## Background

The coronavirus disease 2019 (COVID-19) epidemic has shown the importance of implementing non-pharmaceutical interventions (NPIs), like physical distancing, to contain the spread of the infectious disease and to avoid over-burdening health care systems. As the strain of the virus (SARS-CoV-2) that causes the disease spreads from person to person through infected secretions like saliva or respiratory droplets from the nose or mouth, disease transmission mainly occurs via close social contacts, generally when susceptible individuals breathe in droplets from infected ones, even though transmission through aerosols cannot be ruled out [1–3].

In the absence of a vaccine and effective antiviral treatments, the public health measures introduced to contain the disease were largely NPIs aimed at reducing the number of social contacts, e.g., travel restrictions, school closures, the cancellation of large gatherings, the promotion of physical distancing practices, and, in some countries, nationwide lockdowns. Quantifying changes in social contact patterns in response to these interventions is key to (i) improve our understanding of the determinants of disease transmission, (ii) enable researchers to design more realistic epidemic models, and, ultimately, (iii) assess the effectiveness of public health measures designed to contain the disease and prevent new waves of infection [4, 5]. So far, information about contact patterns under the implemented NPIs has become available only for a limited number of countries in Asia [6], Europe [7–10], Africa [11], and North America [12, 13].

In this study, we leverage new opportunities for data collection, enabled by the digitalization of our lives, to assess changes in social contact patterns on a daily basis and across countries, and to examine their implications for the spread of SARS-CoV-2. We conducted a cross-national online survey, the COVID-19 Health Behavior Survey, for which we recruited participants via targeted Facebook advertisements stratified by sex, age, and subnational region [14, 15]. One goal of the survey was to evaluate the extent to which social contact numbers changed across countries and over time, especially in comparison with those observed prior to the pandemic. We used comparable metrics in our survey, which we conducted in Germany, Italy, the United Kingdom (UK), the United States (US), Spain, France, Belgium, and the Netherlands from March 13 - April 13, 2020, a key period during which the global pandemic was well underway, even though at different stages in different countries. We measured participants’ social contacts, as well as their health behaviors and perceived risk of the coronavirus, on a daily basis and across demographic groups, and quantified the relative impact of the reductions in social contact numbers on the net reproduction number (*R*_*t*_, the average number of secondary infection cases generated by an infectious individual at time *t*) of COVID-19.

## Methods

### Study design

We designed the COVID-19 Health Behavior Survey as a multi-country, cross-sectional, opt-in online survey, to collect key information on people’s health and behavior during the COVID-19 pandemic [14]. Recruitment occurred via targeted Facebook advertisements stratified by sex, age group (18-24 years, 25-44 years, 45-64 years, and 65 years or more), and region of residence, which ensured that a minimum number of respondents could be reached in each stratum [16, 17]. Post-stratification weights by sex, age group, and region, based on nationally representative census data [18, 19], were used in all analyses. Ethical approval for the study was obtained from the Ethics Council of the Max Planck Society, which did not require additional ethics approval for each of the countries included in the study. Participation in this opt-in study was voluntary and open to people who were at least 18 years old and gave their informed consent.

We asked respondents to report the number of individuals with whom they had interacted on the day prior to the survey across different settings, i.e., at home, at school, at work, and in the general community (such as during commuting or leisure activities). We calculated the overall number of daily contacts per person as the sum of contacts in the four different settings, after removing participants who reported more contacts than the 90% quantile of the contact distribution for the respective setting, assuming that they posted high numbers because they were not motivated to take the time to appropriately answer the question. However, our results were robust to the choice of the threshold, as we also considered a fixed cut-off point equal to 29 contacts (used in the most authoritative contact survey in the past [5]), under which contact numbers were generally higher (see Tables S9-S16 in the Additional file 1).

We also asked participants to report on their perceived level of threat related to the coronavirus and on protective behaviors that likely have an impact on disease transmissibility [20], namely, washing their hands more often, wearing a face covering, and avoiding social activities.

### Statistical analysis

We modelled the number of daily contacts reported by participants over the study period using negative binomial regression to account for possible over-dispersion in the contact data and adjusting for respondents’ socio-demographic and behavioral characteristics [21]. The socio-demographic factors that we included as categorical variables were sex, age, region of residence, highest level of education, working status, household size, day of the week of the reported contacts, and being foreign-born. The variable age was also used in an interaction with the week of the reported contacts, to test for different contacts patterns between younger and older adults as time went by. The behavioral factors that we included encompassed dichotomous variables for each protective behavior and a feeling of threat index, based on the simple mean of two different indicators, i.e., the feeling of threat to oneself (much higher for older adults than for the 18-24 group) [14] and the feeling of threat to the family. Interactions between these behavioral factors and the weeks were also included, to account for the time-varying nature of these factors.

### Epidemiological modeling approach

We assessed the consequences of the reduction in social contact numbers for the net reproduction number (*R*_*t*_, i.e., the average number of secondary infections generated by a typical infectious individual at time *t* in a population, either partially or fully susceptible, taking into account the current interventions and the potential spontaneous behavioral change in response to the risk of infection [22–24], with values below one implying that the epidemic is declining and possibly under control). We compared our contact data with those collected prior to the epidemic. For this comparison, we used data from the POLYMOD study for Belgium, Germany, Italy, the Netherlands, and the UK [5], and from the Comes-F study for France [25]. This enabled us to derive age-specific social contact matrices ***C*** compatible with the demographic structure of each country at the beginning of 2020 [26].

We hypothesized (i) that the basic reproduction number *R*_0_ of the infection was consistent with social contact mixing as measured in the pre-COVID period, under the assumption of total absence of immunity in the population and absence of any behavioral change and interventions; (ii) that infection parameters, such as the length of the infectiousness and the infection transmissibility, did not change over time; and (ii) that the initial contact matrices of Italy and the UK could be a reasonable replacement of contact patterns of Spain and the US, because of cultural similarity, after projecting them to the respective demographic structures.

For each week, we obtained a contact matrix 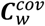 by multiplying the pre-COVID contact matrices ***C***^*pre*^, normalised to the average number of contacts in the population, by the average number of daily contacts by age group that we predicted for each week. More details on the matrix construction can be found in the Supplementary Material. We finally computed the weekly values of 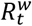 during the study period as 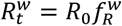, where the weekly reduction factor 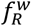 was given by the ratio between the dominant eigenvalue of the weekly contact matrix and that of the pre-COVID matrix, i.e., 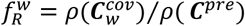.

Our estimates of 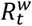 accounted for three sources of uncertainty, namely, (i) the baseline basic reproduction number *R*_0_, which, according to a meta-analysis of the literature, we assumed to be normally distributed with mean 2.6 and standard deviation 0.54, resulting in the 95% confidence interval (CI) (1.54, 3.66) [9, 27]; (ii) the weekly predictions of the daily overall number of contacts, assumed to be normally distributed with mean and standard deviation coming from our predictions; (iii) the pre-COVID contact matrices, which were derived by applying nonparametric bootstrap to the original data. For each source of uncertainty, 5,000 replicates were taken.

## Results

A total of 53,708 individuals completed the questionnaire between March 13 and April 13, 2020. Their distribution by sex, age, and region before and after applying post-stratification weights is shown in Tables S1-S8 in the Supplementary Material.

### Temporal evolution of social contact numbers

Individual social contacts decreased during the study period in all surveyed countries, reaching a plateau at the lowest levels between calendar week 13 (March 23 – 29) and calendar week 14 (March 30 – April 5) (Figure 1). Evidence from the UK and the US showed that the overall numbers of daily social contacts declined considerably after governments issued physical distancing guidelines, which mainly occurred during week 11 (March 9 – 15), rather than after more stringent lockdown measures were announced. Comparing trends before and after the announcement of physical distancing measures, both occurring between March 10 and March 19 [28, 29], we estimated that the average number of daily contacts (with 95% CI) decreased from 5.70 (4.83, 6.57) in midweek 11 to 3.43 (2.67, 4.19) in midweek 12, representing for the US a decrease of 40%, and from 6.06 (4.77, 7.35) to 4.04 (2.63, 5.45) in the UK, representing a reduction of 33%. In midweek 14, after the implementation of lockdown measures, contact numbers decreased even further in the UK to 2.68 (2.05, 3.31), but not in the US, where contact numbers had essentially plateaued since the beginning of week 13. In the other countries, where physical distancing guidelines were introduced before the beginning of our study, we found that the average number of contacts had already declined to their lowest levels in midweek 13, ranging from 2.31 (1.93, 2.69) in France to 3.87 (3.06, 4.68) in Germany.

**Figure 1.**
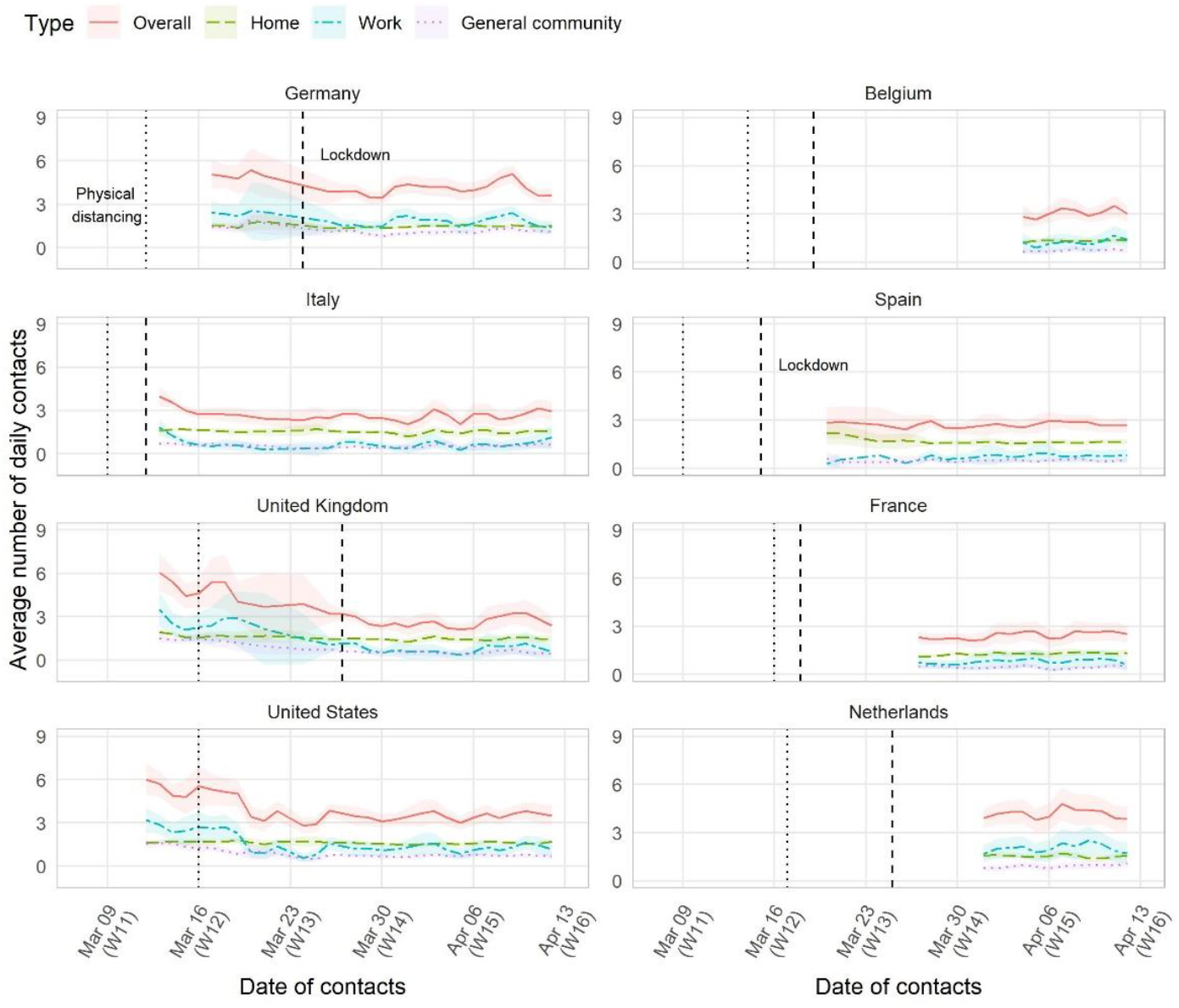
Mean overall number of daily social contacts (with 95% CI) smoothed by a simple two-day moving average, by country and study day, March-April 2020. The dotted line corresponds to the date in which the physical distancing guidelines were introduced at the national level; the dashed line corresponds to the date in which the lockdown, regardless of being full or partial, was ordered.

Reductions in social contacts differed between those countries that eventually applied a full lockdown (people were required to stay at home and not allowed to go out if not necessary), such as Belgium, France, Italy, Spain, and the UK, and those that opted for a partial lockdown (a stricter version of the physical distancing guidelines was imposed, but people had the freedom to go out), such as Germany and the Netherlands (Table 1). For the latter two countries, we estimated that each person had more than four contacts per day in week 15, a substantially higher value than those estimated for countries with full lockdown (up to three per day). The comparison between the pre-pandemic contact data [5, 25] and those in week 15 showed that the largest reduction had occurred in Italy, at 83%, though substantial declines also occurred in the other countries, ranging from 49% in Germany to between 71% and 76% in Belgium, France, the Netherlands, and the UK

**Table 1.**
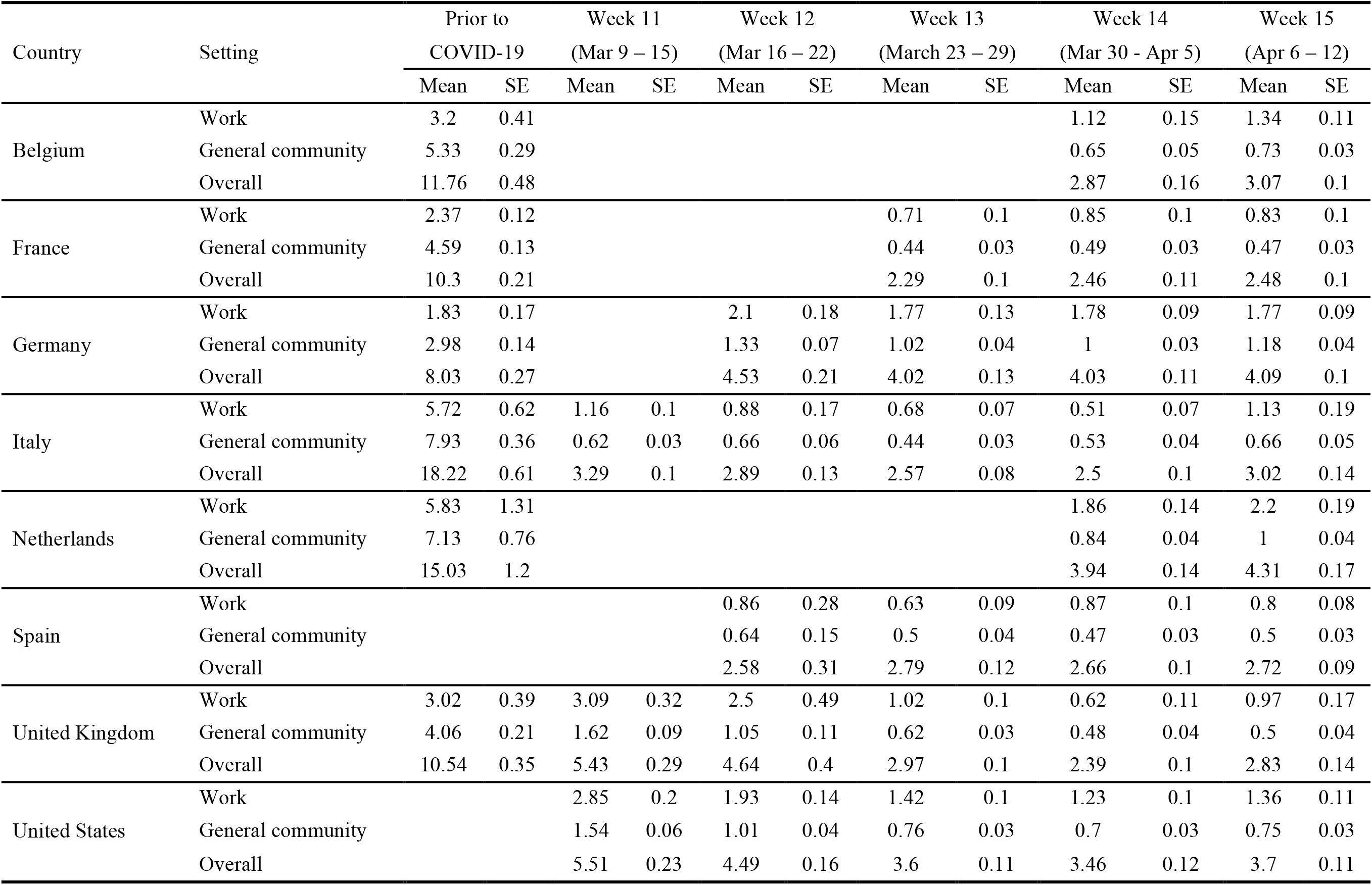
Model-predicted mean number (with SE) of daily contacts per person compared with pre-pandemic model predictions, by country, setting, and week, March-April 2020.

| Country | Setting | Prior to<br>COVID-19 |  | Week 11<br>(Mar 9 – 15) |  | Week 12<br>(Mar 16 – 22) |  | Week 13<br>(March 23 – 29) |  | Week 14<br>(Mar 30 - Apr 5) |  | Week 15<br>(Apr 6 – 12) |  |
| --- | --- | --- | --- | --- | --- | --- | --- | --- | --- | --- | --- | --- | --- |
|  |  | Mean | SE | Mean | SE | Mean | SE | Mean | SE | Mean | SE | Mean | SE |
| Belgium | Work | 3.2 | 0.41 |  |  |  |  |  |  | 1.12 | 0.15 | 1.34 | 0.11 |
|  | General community | 5.33 | 0.29 |  |  |  |  |  |  | 0.65 | 0.05 | 0.73 | 0.03 |
|  | Overall | 11.76 | 0.48 |  |  |  |  |  |  | 2.87 | 0.16 | 3.07 | 0.1 |
| France | Work | 2.37 | 0.12 |  |  |  |  | 0.71 | 0.1 | 0.85 | 0.1 | 0.83 | 0.1 |
|  | General community | 4.59 | 0.13 |  |  |  |  | 0.44 | 0.03 | 0.49 | 0.03 | 0.47 | 0.03 |
|  | Overall | 10.3 | 0.21 |  |  |  |  | 2.29 | 0.1 | 2.46 | 0.11 | 2.48 | 0.1 |
| Germany | Work | 1.83 | 0.17 |  |  | 2.1 | 0.18 | 1.77 | 0.13 | 1.78 | 0.09 | 1.77 | 0.09 |
|  | General community | 2.98 | 0.14 |  |  | 1.33 | 0.07 | 1.02 | 0.04 | 1 | 0.03 | 1.18 | 0.04 |
|  | Overall | 8.03 | 0.27 |  |  | 4.53 | 0.21 | 4.02 | 0.13 | 4.03 | 0.11 | 4.09 | 0.1 |
| Italy | Work | 5.72 | 0.62 | 1.16 | 0.1 | 0.88 | 0.17 | 0.68 | 0.07 | 0.51 | 0.07 | 1.13 | 0.19 |
|  | General community | 7.93 | 0.36 | 0.62 | 0.03 | 0.66 | 0.06 | 0.44 | 0.03 | 0.53 | 0.04 | 0.66 | 0.05 |
|  | Overall | 18.22 | 0.61 | 3.29 | 0.1 | 2.89 | 0.13 | 2.57 | 0.08 | 2.5 | 0.1 | 3.02 | 0.14 |
| Netherlands | Work | 5.83 | 1.31 |  |  |  |  |  |  | 1.86 | 0.14 | 2.2 | 0.19 |
|  | General community | 7.13 | 0.76 |  |  |  |  |  |  | 0.84 | 0.04 | 1 | 0.04 |
|  | Overall | 15.03 | 1.2 |  |  |  |  |  |  | 3.94 | 0.14 | 4.31 | 0.17 |
| Spain | Work |  |  |  |  | 0.86 | 0.28 | 0.63 | 0.09 | 0.87 | 0.1 | 0.8 | 0.08 |
|  | General community |  |  |  |  | 0.64 | 0.15 | 0.5 | 0.04 | 0.47 | 0.03 | 0.5 | 0.03 |
|  | Overall |  |  |  |  | 2.58 | 0.31 | 2.79 | 0.12 | 2.66 | 0.1 | 2.72 | 0.09 |
| United Kingdom | Work | 3.02 | 0.39 | 3.09 | 0.32 | 2.5 | 0.49 | 1.02 | 0.1 | 0.62 | 0.11 | 0.97 | 0.17 |
|  | General community | 4.06 | 0.21 | 1.62 | 0.09 | 1.05 | 0.11 | 0.62 | 0.03 | 0.48 | 0.04 | 0.5 | 0.04 |
|  | Overall | 10.54 | 0.35 | 5.43 | 0.29 | 4.64 | 0.4 | 2.97 | 0.1 | 2.39 | 0.1 | 2.83 | 0.14 |
| United States | Work |  |  | 2.85 | 0.2 | 1.93 | 0.14 | 1.42 | 0.1 | 1.23 | 0.1 | 1.36 | 0.11 |
|  | General community |  |  | 1.54 | 0.06 | 1.01 | 0.04 | 0.76 | 0.03 | 0.7 | 0.03 | 0.75 | 0.03 |
|  | Overall |  |  | 5.51 | 0.23 | 4.49 | 0.16 | 3.6 | 0.11 | 3.46 | 0.12 | 3.7 | 0.11 |

### Temporal evolution of protecting behavior

Reductions in social contacts were accompanied by an increased use of a face covering in all countries, except for Belgium and the Netherlands (stable at 10%) [14]. The largest increases were observed in Italy (from 40% in week 11 to 80% in week 15) and in the US (from 10% in week 11 to 60% in week 15), while in the UK it reached 20% by the same date (Figure 2). Between 80% and 90% of participants in every country reported avoiding social activities, with evidence of a sharp increase over time in the UK and, to a lesser extent, the US. Finally, little to no variation was observed for handwashing, for which we observed a prevalence level of around 90% that was stable over time; and the feeling of threat, which ranged between 50% and 60% in all countries.

**Figure 2.**
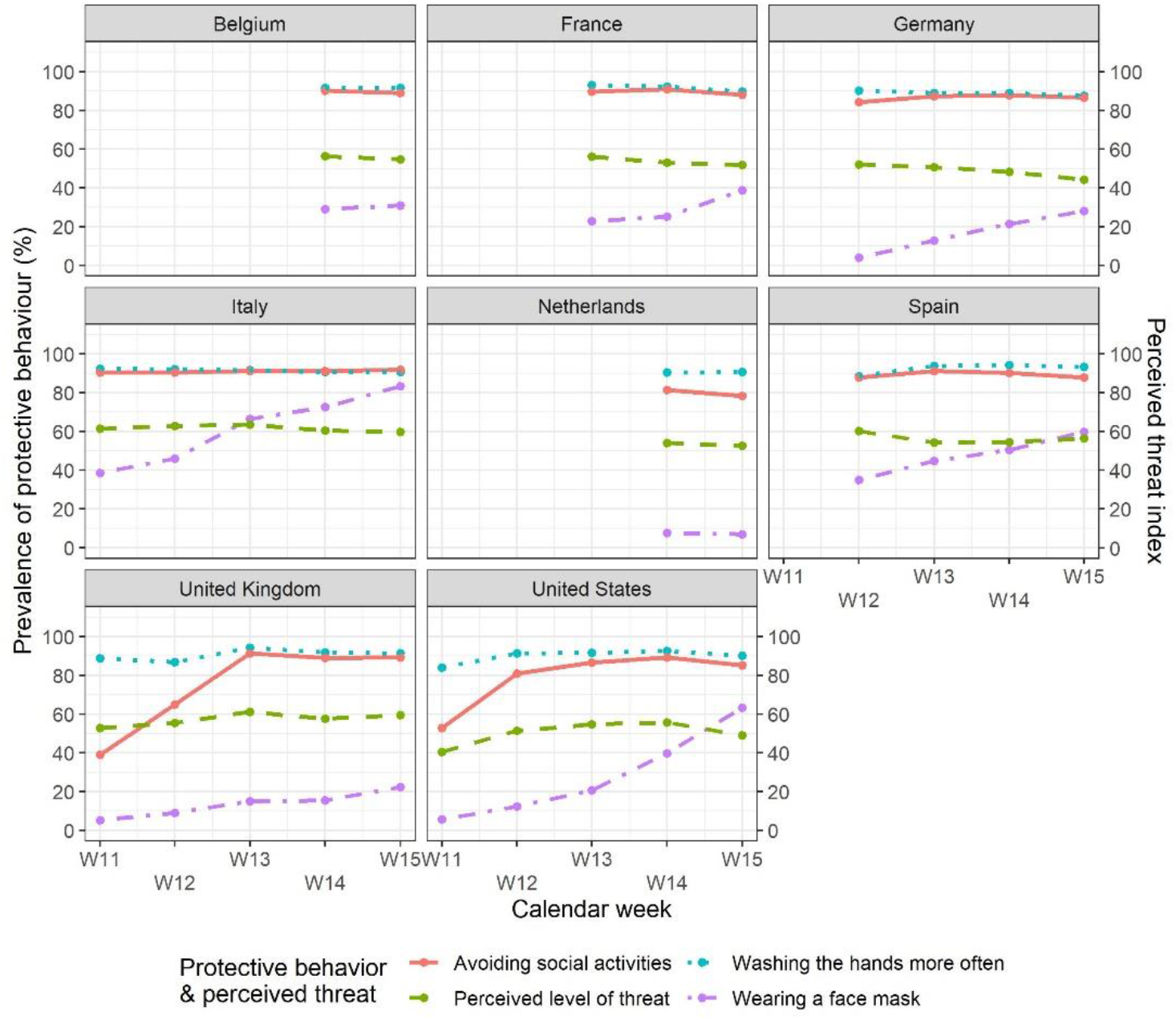
Prevalence of participants’ uptake of different behavioral measures to protect themselves from the coronavirus and perceived level of threat (averaging the perceived threat to oneself with that to the family, after rescaling to a 0-100 scale), by country and week.

### Differences in contact numbers by age group

The evolution of the model-predicted overall daily contact numbers by week showed a decreasing variation in social contact numbers that was uniform across age groups. Higher social contact numbers were estimated for people aged 18-24 years, while those aged 65+ years usually reported the lowest numbers from the beginning of the study period. The reduction in the overall contact numbers was especially evident when comparing survey data for week 15 to pre-COVID data (Figure 3). Similar results obtained under an alternative cut-off threshold for outliers are shown in Figure S9 in the Supplementary Material.

**Figure 3.**
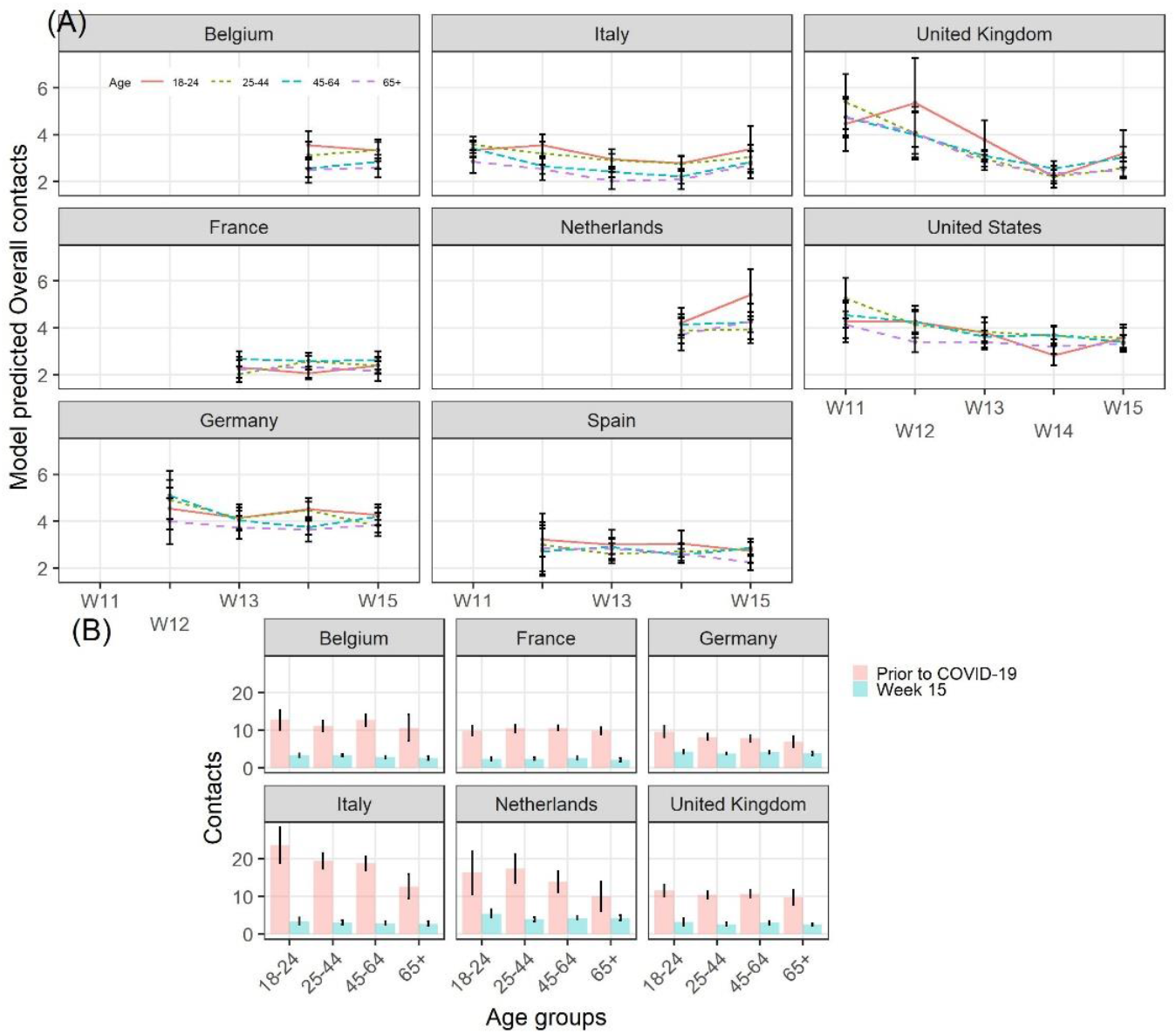
Model-predicted overall daily number of social contacts, by age group, country, and week, March-April 2020. (A). Comparison of model-predicted overall contact numbers between the pre-COVID period and calendar week 15, by country (B).

### Assessment of the net reproduction number

Finally, we found that the weekly net reproduction number 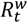 generally decreased from the baseline value of the basic reproduction number *R*_0_ under social age mixing in the period prior to COVID-19, albeit with large heterogeneity among countries (Figure 4). In Italy, Spain, France, and Belgium, the reduction in contact numbers led to a decrease between 70% and 80% of the baseline *R*_0_, and hence to a 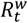 below one, as early as in week 11, while in the UK, evidence of the 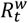 below one did not appear until week 14. In the Netherlands and the US, the 95% CI of the 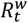 always included the value of one, following reductions of around 60-70% in both contact numbers and 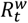. Finally, in Germany, we observed that the decline in the overall contact numbers led to a rather small decrease in the 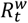, which reached its minimum value, 1.45 (0.86–2.07), in week 15.

**Figure 4.**
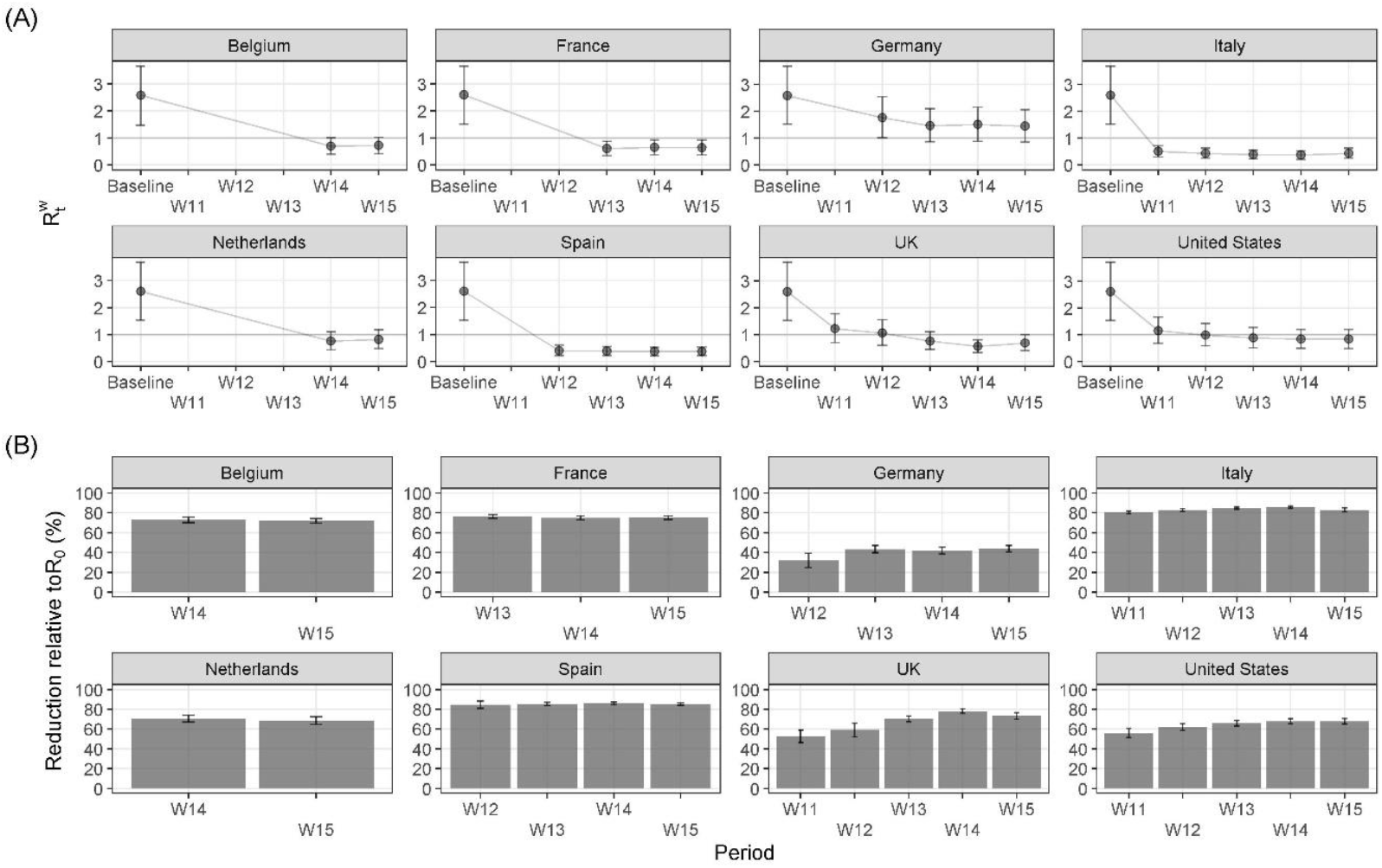
Absolute change in the weekly net reproduction number 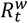 with respect to the basic reproduction number *R*_0_ at baseline (A) and percent reduction with respect to *R*_0_ (B), by country and week. The 95% CIs are based on 5,000 replicates.

Under the alternative threshold, we found that the higher number of contacts entailed lower reductions in both social contacts and in the 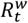, which was below one for Italy and Spain, and equal to one for Belgium, France, Netherlands, and, from week 13, the UK and the US (Figure S12 in the Supplementary Material).

## Discussion

This paper provided comparative estimates of the daily number of social contacts, overall and by setting of contact, that people had during the early phase of the COVID-19 pandemic, between March 12 (week 11) and April 12 (week 15), in Belgium, France, Germany, Italy, the Netherlands, Spain, the UK, and the US, disaggregated by week. Compared to the pre-COVID period, we estimated a large decrease in the overall contact numbers, which was however not uniform across all countries. Such reductions were smaller than that of 86% observed in Wuhan (China) at the beginning of the epidemic [6], as they ranged from 49% in Germany to 83% in Italy. Our estimates were consistent with those from other independent studies conducted at the same time in Europe and the US [7–10, 13, 30]. The unique aspect of our study is that it is the only one to use a consistent approach to collect comparative data across several countries and over an extended period of time.

Our findings suggest that the countries with the highest numbers of social contacts were those that implemented partial lockdowns (or stricter versions of the physical distancing guidelines), whereas the countries that implemented full lockdown measures experienced much lower contact numbers. Evidence from the UK and the US showed that the numbers of contacts declined sharply after governments issued physical distancing guidelines, even before the introduction of the lockdown measures. This pattern was also suggested by mobile phone data made available by Google, illustrating the change in human mobility compared to January and February 2020 [31, 32].

We also estimated the impact of reductions in social contacts on the weekly net reproduction number 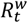. An 80% reduction in contacts was associated with an equivalent reduction in th 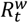, which was compatible with values below one in Italy, Spain, France, Belgium, and the UK, and equal to one in the Netherlands and the US. The exception was Germany, where the 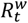 estimate was always above one. Indeed, independent estimates of *Rt* at the beginning of April in Germany, i.e., 1.1 with 95% CI 0.9–1.4, were consistent with those presented in this paper [33, 34]. These findings can be explained by the relatively low social contact numbers in Germany during the time prior to COVID-19 (eight contacts per day, on average) and by the smaller reductions in social contact numbers during the study period (between 40% and 50%). Nonetheless, these results should not be interpreted as evidence that the infection spread was exceptionally high in Germany despite physical distancing interventions. Changes in the *R*_*t*_ are brought about not only by changes in social contact patterns, but also in infection transmissibility, which might be affected by several additional factors, such as travel restrictions, keeping the distance between people while having conversations in public spaces, increased hand washing or sanitizing, and the use of face masks [29, 35, 36], which could be considered only partially in our calculations.

The study has limitations that should be discussed considering the findings. First, we collected data using an opt-in sample of Facebook users. Such non-probabilistic samples are somewhat less accurate than probability samples, but with the appropriate statistical adjustments such as those that we made, they offer a good approximation to results that can be obtained from probabilistic samples. Furthermore, such samples can be collected rapidly and therefore can provide us with timely data that could not possibly be collected otherwise during a pandemic. Finally, our data cover a large population at minimal costs compared to more traditional surveys [15].

Second, as survey respondents were recruited through targeted online advertisements, this approach may potentially lead to self-selection bias, which, with online surveys on coronavirus-related behavioral outcomes, has been linked to the risk of underestimating the share of population complying with specific measures [37]. Indeed, our participants might differ to some extent from the general population in terms of their sociability patterns (due to their Facebook use) and their concerns about health-related issues (as they opted in to participate in the survey). We tackled this issue by stratifying our advertising campaigns and constructing post-stratification weights by factors that we believe are linked to both survey participation and the outcomes of interest of the survey [38, 39], and by ensuring that only respondents who were shown the Facebook advertisement and clicked on the link could participate in the study (thereby avoiding spill over biases in the sample that we cannot adjust for). However, post-stratification weights cannot correct entirely for different behaviors or social inequalities among Facebook users since such variables cannot be included in the marketing campaign stratification.

Third, several survey participants reported extremely high contact numbers, perhaps because they had little motivation to provide accurate counts. This is a general problem in social contact surveys. Typically, social contact surveys either include a maximum number of contacts that respondents can report, or researchers assess ex-post the threshold level for the right tail of the contact distribution of contacts. We followed the latter approach for the removal of respondents reporting implausible values. However, we also found that our results are robust to a choice of cut-off values consistent with those used in standard surveys in the field. Future contact surveys may consider including attention checks items to reduce potential issues with outliers.

## Conclusions

In conclusion, this paper provides a large international comparable set of statistics on social contact patterns during the COVID-19 pandemic, disaggregated by country, setting, and week. As these estimates offer a more grounded alternative to the theoretical assumptions that have so far been used [35, 40, 41], we believe that the scientific community can draw on this information for developing more realistic epidemic models of COVID-19.

## Data Availability

All the primary data needed to evaluate the conclusions in the paper are currently stored in a secure and protected server at the Max Planck Institute for Demographic Research, and are not publicly available in order to comply with data protection regulations in Germany, and to preserve the privacy of study participants. Replication data and R/Stata code for all our findings will be made available in the future once the appropriate privacy measures have been implemented to avoid identification of individuals in the microdata.

## Notes

### Competing Interest Statement

The authors have declared no competing interest.

### Funding Statement

This study was funded through the support of the Max Planck Institute for Demographic Research, which is part of the Max Planck Society.

### Author Declarations

Ethical approval for the study was obtained from the Ethics Council of the Max Planck Society, which did not require additional ethics approval for each of the countries included in the study. Participation in this opt-in study was voluntary and open to people who were at least 18 years old and gave their informed consent.

### Summary of Updates

Updated methodology to compute social contact matrices based on new estimates of contact numbers by age group. Updated model for estimating contacts, added variables on health behavior and perceived level of threat. Updated sections on epidemiological analysis (methods and results). Added sensitivity analysis on threshold at 30 contacts per day. Author affiliation updated. Figure 1 revised, based on observed data, rather than on predictions. Figure 2 added based on data on health behavior and perceived level of threat. Figure 3 revised, comparing pre-pandemic estimates by age with those from week 15. Figure 4 revised, based on updated epidemiological analysis.

## References

1. Bahl P, Doolan C, de Silva C, Chughtai AA, Bourouiba L, MacIntyre CR. Airborne or Droplet Precautions for Health Workers Treating Coronavirus Disease 2019? J Infect Dis. 2020.

2. Chu DK, Akl EA, Duda S, Solo K, Yaacoub S, Schünemann HJ, et al. Physical distancing, face masks, and eye protection to prevent person-to-person transmission of SARS-CoV-2 and COVID-19: a systematic review and meta-analysis. The Lancet. 2020;395:1973–87.

3. WHO.Transmission of SARS-CoV-2: implications for infection prevention precautions. 2020. https://www.who.int/news-room/commentaries/detail/transmission-of-sars-cov-2-implications-for-infection-prevention-precautions. Accessed 24 Aug 2020.

4. Wallinga J, Teunis P, Kretzschmar M. Using Data on Social Contacts to Estimate Age-specific Transmission Parameters for Respiratory-spread Infectious Agents. Am J Epidemiol. 2006;164:936–44.

5. Mossong J, Hens N, Jit M, Beutels P, Auranen K, Mikolajczyk R, et al. Social Contacts and Mixing Patterns Relevant to the Spread of Infectious Diseases. PLoS Med. 2008;5:e74.

6. Zhang J, Litvinova M, Liang Y, Wang Y, Wang W, Zhao S, et al. Changes in contact patterns shape the dynamics of the COVID-19 outbreak in China. Science. 2020;368:1481–6.

7. Backer JA, Mollema L, Vos ERA, Klinkenberg D, Klis FRM van der, Melker HE de, et al. The impact of physical distancing measures against COVID-19 transmission on contacts and mixing patterns in the Netherlands: repeated cross-sectional surveys in 2016/2017, April 2020 and June 2020. medRxiv. 2020;:2020.05.18.20101501.

8. Coletti P, Wambua J, Gimma A, Willem L, Vercruysse S, Vanhoutte B, et al. CoMix: comparing mixing patterns in the Belgian population during and after lockdown. Sci Rep. 2020;10:21885.

9. Jarvis CI, Van Zandvoort K, Gimma A, Prem K, CMMID COVID-19 working group,Klepac P, et al. Quantifying the impact of physical distance measures on the transmission of COVID-19 in the UK. BMC Med. 2020;18:124.

10. Latsuzbaia A, Herold M, Bertemes J-P, Mossong J. Evolving social contact patterns during the COVID-19 crisis in Luxembourg. PLoS ONE. 2020;15:e0237128.

11. Quaife M, van Zandvoort K, Gimma A, Shah K, McCreesh N, Prem K, et al. The impact of COVID-19 control measures on social contacts and transmission in Kenyan informal settlements. BMC Med. 2020;18:316.

12. Dorélien AM, Simon A, Hagge S, Call KT, Enns E, Kulasingam S. Minnesota Social Contacts and Mixing Patterns Survey with Implications for Modelling of Infectious Disease Transmission and Control. Surv Pract. 2020;13:13669.

13. Feehan D, Mahmud A. Quantifying population contact patterns in the United States during the COVID-19 pandemic. medRxiv. 2020;:2020.04.13.20064014.

14. Perrotta D, Grow A, Rampazzo F, Cimentada J, Del Fava E, Gil-Clavel S, et al. Behaviors and attitudes in response to the COVID-19 pandemic: Insights from a cross-national Facebook survey. medRxiv. 2020;:2020.05.09.20096388.

15. Grow A, Perrotta D, Fava ED, Cimentada J, Rampazzo F, Gil-Clavel S, et al. Addressing Public Health Emergencies via Facebook Surveys: Advantages, Challenges, and Practical Considerations. J Med Internet Res. 2020;22:e20653.

16. Pötzschke S, Braun M. Migrant Sampling Using Facebook Advertisements: A Case Study of Polish Migrants in Four European Countries. Soc Sci Comput Rev. 2016;35:633–53.

17. Zhang B, Mildenberger M, Howe PD, Marlon J, Rosenthal SA, Leiserowitz A. Quota sampling using Facebook advertisements. Polit Sci Res Methods. 2018;:1–7.

18. Eurostat, Statistical Office of the European Union. Eurostat regional yearbook: 2019 edition. 2019. http://publications.europa.eu/publication/manifestation_identifier/PUB_KSHA19001ENN. Accessed 15 May 2020.

19. US Census Bureau. 2018 Population Estimates by Age, Sex, Race and Hispanic Origin. The United States Census Bureau. https://www.census.gov/newsroom/press-kits/2019/detailed-estimates.html. Accessed 15 May 2020.

20. Teslya A, Pham TM, Godijk NG, Kretzschmar ME, Bootsma MCJ, Rozhnova G. Impact of self-imposed prevention measures and short-term government-imposed social distancing on mitigating and delaying a COVID-19 epidemic: A modelling study. PLoS Med. 2020;17:e1003166.

21. Hens N, Ayele GM, Goeyvaerts N, Aerts M, Mossong J, Edmunds JW, et al. Estimating the impact of school closure on social mixing behaviour and the transmission of close contact infections in eight European countries. BMC Infect Dis. 2009;9:187.

22. Liu Q-H, Ajelli M, Aleta A, Merler S, Moreno Y, Vespignani A. Measurability of the epidemic reproduction number in data-driven contact networks. Proc Natl Acad Sci. 2018;115:12680–5.

23. The Royal Society. Reproduction number (R) and growth rate (r) of the COVID-19 epidemic in the UK. 2020. https://royalsociety.org/news/2020/09/set-c-covid-r-rate/.

24. Zhang J, Litvinova M, Wang W, Wang Y, Deng X, Chen X, et al. Evolving epidemiology and transmission dynamics of coronavirus disease 2019 outside Hubei province, China: a descriptive and modelling study. Lancet Infect Dis. 2020;20:793–802.

25. Béraud G, Kazmercziak S, Beutels P, Levy-Bruhl D, Lenne X, Mielcarek N, et al. The French Connection: The First Large Population-Based Contact Survey in France Relevant for the Spread of Infectious Diseases. PLoS ONE. 2015;10:e0133203.

26. Arregui S, Aleta A, Sanz J, Moreno Y. Projecting social contact matrices to different demographic structures. PLoS Comput Biol. 2018;14:e1006638.

27. Anderson RM, Heesterbeek H, Klinkenberg D, Hollingsworth TD. How will country-based mitigation measures influence the course of the COVID-19 epidemic? The Lancet. 2020;395:931–4.

28. Mervosh S, Lu D, Swales V. See Which States and Cities Have Told Residents to Stay at Home. The New York Times. 2020. https://www.nytimes.com/interactive/2020/us/coronavirus-stay-at-home-order.html. Accessed 25 Sep 2020.

29. Flaxman S, Mishra S, Gandy A, Unwin HJT, Mellan TA, Coupland H, et al. Estimating the effects of non-pharmaceutical interventions on COVID-19 in Europe. Nature. 2020;584:257–61.

30. Sypsa V, Roussos S, Paraskevis D, Lytras T, Tsiodras S, Hatzakis A. Modelling the SARS-CoV-2 first epidemic wave in Greece: social contact patterns for impact assessment and an exit strategy from social distancing measures. medRxiv. 2020;:2020.05.27.20114017.

31. Google LLC. Google COVID-19 Community Mobility Reports. https://www.google.com/covid19/mobility/. Accessed 22 Jun 2020.

32. Basellini U, Alburez-Gutierrez D, Del Fava E, Perrotta D, Bonetti M, Camarda CG, et al. Linking excess mortality to Google mobility data during the COVID-19 pandemic in England and Wales. SocArXiv. 2020. doi:10.31235/osf.io/75d6m.

33. RKI. RKI - Coronavirus SARS-CoV-2 - Situation report - 9 Apr 2020. Situation Report. Robert Koch Institute; 2020. https://www.rki.de/DE/Content/InfAZ/N/Neuartiges_Coronavirus/Situationsberichte/2020-04-09-en.pdf?blob=publicationFile,. Accessed 15 May 2020.

34. Barbarossa MV, Fuhrmann J, Meinke JH, Krieg S, Varma HV, Castelletti N, et al. Modeling the spread of COVID-19 in Germany: Early assessment and possible scenarios. PLoS ONE. 2020;15:e0238559.

35. Dehning J, Zierenberg J, Spitzner FP, Wibral M, Neto JP, Wilczek M, et al. Inferring change points in the spread of COVID-19 reveals the effectiveness of interventions. Science. 2020;369.

36. Chinazzi M, Davis JT, Ajelli M, Gioannini C, Litvinova M, Merler S, et al. The effect of travel restrictions on the spread of the 2019 novel coronavirus (COVID-19) outbreak. Science. 2020. doi:10.1126/science.aba9757.

37. Schaurer I, Weiß B. Investigating selection bias of online surveys on coronavirus-related behavioral outcomes. Surv Res Methods. 2020;14:103–8.

38. Keiding N, Louis TA. Perils and potentials of self-selected entry to epidemiological studies and surveys. J R Stat Soc Ser A Stat Soc. 2016;179:319–76.

39. Mercer AW, Kreuter F, Keeter S, Stuart EA. Theory and Practice in Nonprobability Surveys Parallels between Causal Inference and Survey Inference. Public Opin Q. 2017;81:250–71.

40. Gatto M, Bertuzzo E, Mari L, Miccoli S, Carraro L, Casagrandi R, et al. Spread and dynamics of the COVID-19 epidemic in Italy: Effects of emergency containment measures. Proc Natl Acad Sci. 2020;117:10484–91.

41. Salje H, Kiem CT, Lefrancq N, Courtejoie N, Bosetti P, Paireau J, et al. Estimating the burden of SARS-CoV-2 in France. Science. 2020;369:208–11.

