## Supplementary Material for "The differential impact of physical distancing strategies on social contacts relevant for the spread of COVID-19: Evidence from a multi-country survey"

### Additional methods

#### Survey design

The data used in the analysis came from the CHBS, an online survey carried out in multiple countries (and multiple languages) to collect key information about people's health and behavior in a time of growing uncertainty due to the COVID-19 pandemic [1]. Participation was voluntary and anonymous, and open to people who were at least 18 years old. Recruitment occurred through targeted advertisements implemented through the Facebook Ads Manager (FAM), which is a tool that can be used to quickly reach large numbers of survey participants across several countries. The online survey was stratified by age groups, sex, and region of residence (largely following the NUTS-1 classification in Europe and the census regions in the United States, both aggregated into larger macro-regions) using the FAM to ensure that a minimum number of respondents was reached in all strata.

The questionnaire was divided into multiple sections covering different areas of interest: socio-demographic indicators (age, gender, country of birth, region of residence, level of education, and household size); health indicators (symptoms experienced in the previous seven days, among others); opinions and behaviors (preventive measures taken and disruption to daily routine, among others); and social contact data. For the purposes of validation and comparability, the questionnaire included standard questions taken from relevant sources, such as the European Social Survey for socio-demographic questions [2], and an Ipsos poll for questions related to public opinion on the COVID-19 outbreak [3].

Informed consent was obtained from all participants, who had to be 18 years or older, enabling the collection, storage, and processing of their answers. Participants' data was treated anonymously. Ethical approval for the study was obtained from the Ethics Council of the Max Planck Society.

#### Recruitment Strategy

Marketers use the FAM to advertise their products or businesses. The FAM enables advertisers to create advertising campaigns that can have various goals, such as creating salience for a given service or product among Facebook users, or generating traffic to an external website. Following previous examples of Facebook surveys [4–6], we used the FAM to run campaigns to recruit respondents for our study. The countries included in the study, selected because of the number of COVID-19 cases they were experiencing, were Germany, Italy, Spain, the UK, the US, France, Belgium, and the Netherlands. The FAM allowed us to perform a quota sampling by targeting user groups based on demographic characteristics, such as sex, age, and region of residence. In this way, not only we could ensure a minimum number of participants in each combination of strata, but we were also able to control for variables possibly associated with both survey participation and the survey outcomes [7, 8].

At the beginning of March, prior to the start of the actual survey, we carried out separate pilots in Italy, the

UK, and the US to test the FAM and the performance of our ads, allowing the study to officially begin on March 13. Figure S1 shows the question that prompted survey participants to report the information on their social contacts in the English-language version of the questionnaire.

Due to technical problems on the Facebook platform for some countries, very few respondents accessed the survey on certain days between March 20 and March 25. We thus excluded from the analysis those days with less than ten respondents. Although this issue might have increased the uncertainty in the model predictions (especially the daily ones), we believe that the analysis by week is robust and less affected by this issue.

#### Poststratification

Since online surveys are not random samples of the population, we adjusted our survey data using a poststratification weighting approach. In particular, given that our aim was to achieve a nationally representative sample, we stratified the survey by age group, sex, and region of residence, which are all important variables that are related to differences in people's responses to the pandemic, as well as to survey participation. In order to create poststratification weights, we divided the true population proportion in each stratum, obtained from nationally representative data available through Eurostat (2019) [2] and the US census (2018) [9], by the sample proportion from the same stratum in our survey. These poststratification weights were used in all the statistical analyses presented in this paper.

#### Social contact data

We defined social contacts, which are the focus of this paper, as any interaction involving either physical contact (such as a handshake or a hug) or a conversation of three or more words in the physical presence of another person. This definition is consistent with the definition employed in past social contact surveys, which we used as the baseline for comparison in this work [10, 11]. In more detail, we asked respondents to report the number of individuals with whom they interacted on the day before the survey in different settings: i.e., at home, at school or college, at work, and in the general community (such as during commuting or leisure activities); while making it clear that the respondent should not report multiple interactions with the same person in different settings. However, unlike previous contact surveys, we did not ask about the characteristics of the contacted individuals (e.g., age and sex) to avoid overburdening respondents, given the nature of the online survey. The text describing the social contact question is shown in Figure S1.

#### Health behaviors and perceived risk

We asked respondents to rate the level of perceived threat they thought the COVID-19 posed to different levels of society, namely, (i) to themselves, (ii) to their family, (iii) to their local community, (iv) to their country, and (v) to the world, using a 5-point Likert scale. For the sake of our analysis, we rescaled participants' responses to the range 0 – 1, where zero indicated "very low threat" and one "very high threat". To account for a feeling of threat that might have affected social contact patterns of respondents, we combined in one variable the perceived level of threat to themselves and to their families, taking their simple mean. These two indicators were found to have a good internal consistency, having a Cronbach's alpha of 0.84.

Moreover, we asked respondents which health behaviors, in the form of specific actions, they took up to protect themselves from the virus. Among the listed actions, coded as dummy variables, we asked whether respondents avoided shaking hands, social activities (e.g., meeting friends), crowded places (e.g., restaurants, cinemas, gym, or playground), travelling by public transportation (e.g., by bus, tram, subway, or train) or by taxi, whether they stockpiled medicines or food, whether they used a face mask, and whether they washed their hands or used a sanitizing hand gel more often. For our analysis, we picked three of these actions, which might have an impact on the transmissibility of the virus, namely, avoiding social activities, wearing a face mask, and washing the hands more often [12, 13].

### Epidemiological analysis

The net reproduction number $R_t$ can be estimated as the dominant eigenvalue, denoted as $\rho$, of the next generation matrix $\boldsymbol{N}$, i.e., $R_t = \rho(\boldsymbol{N})$, where $\boldsymbol{N}$ provides information on the numbers of newly infected individuals by age group at a given time [14]. Under the "social contact hypothesis", $\boldsymbol{N} = Dq\boldsymbol{C}$, where $\boldsymbol{C}$ is the matrix containing the average number of contacts between age groups, $q$ is the disease transmissibility parameter, and $D$ is the length of the infectiousness period [15]. We use the proportional relationship between $\boldsymbol{N}$ and $\boldsymbol{C}$, i.e., $\boldsymbol{N} = \frac{R_t}{\rho(\boldsymbol{C})}\boldsymbol{C}$, to assess changes in $R_t$ due to changes in social contact numbers [16–19].

As we did not collect information on the age of the individuals encountered by the study participants, we derived the contact matrices $\boldsymbol{C}$ for each study week using the following procedure: (i) for each country, based on the data collected in the POLYMOD study between 2005 and 2006 [10], and in the Comes-F studies in 2012 [11], we constructed pre-COVID age-specific social contact matrices $\boldsymbol{C}^{pre}$, using the same age groups of the survey (18 – 24 years, 25 – 44 years, 45 – 64 years, and 65 years or more), which contained the average number of contacts between participants in the $i$th age group and their contactees in the $j$th age group; (ii) we projected the matrices $\boldsymbol{C}^{pre}$ to the population structure of each country in 2020 [20]; (ii) to remove differences in the contact levels between countries, we normalized each matrix $\boldsymbol{C}^{pre}$ to the average number of contacts at the population level, so that the mean number of contacts of the matrix $\boldsymbol{C}^{pre+norm}$ was equal to one; (iii) we multiplied the normalized matrix $\boldsymbol{C}^{pre+norm}$ by the average number of daily contacts that we predicted for each group and study week, thus obtaining the weekly COVID matrices $\boldsymbol{C}_w^{cov}$.

As we did not collect data for people younger than 18 years old, we computed their average number of daily contacts per week in the following way. To reflect school closure during the survey period, we excluded school contacts from the contact data prior to COVID-19, considering only the overlapping age groups between the two types of matrices. Then, for each country, we computed a weekly overall scaling factor, and we obtained the average number of contacts for individuals aged less than 18 by multiplying their average number of pre-COVID-19 contacts in the population for such factor.

To account for the uncertainty in the pre-COVID contact data, we applied a non-parametric bootstrap procedure to the original data, resampling with replacement the participants, proportionally to the population age distribution, and assigning them to their original contact data.

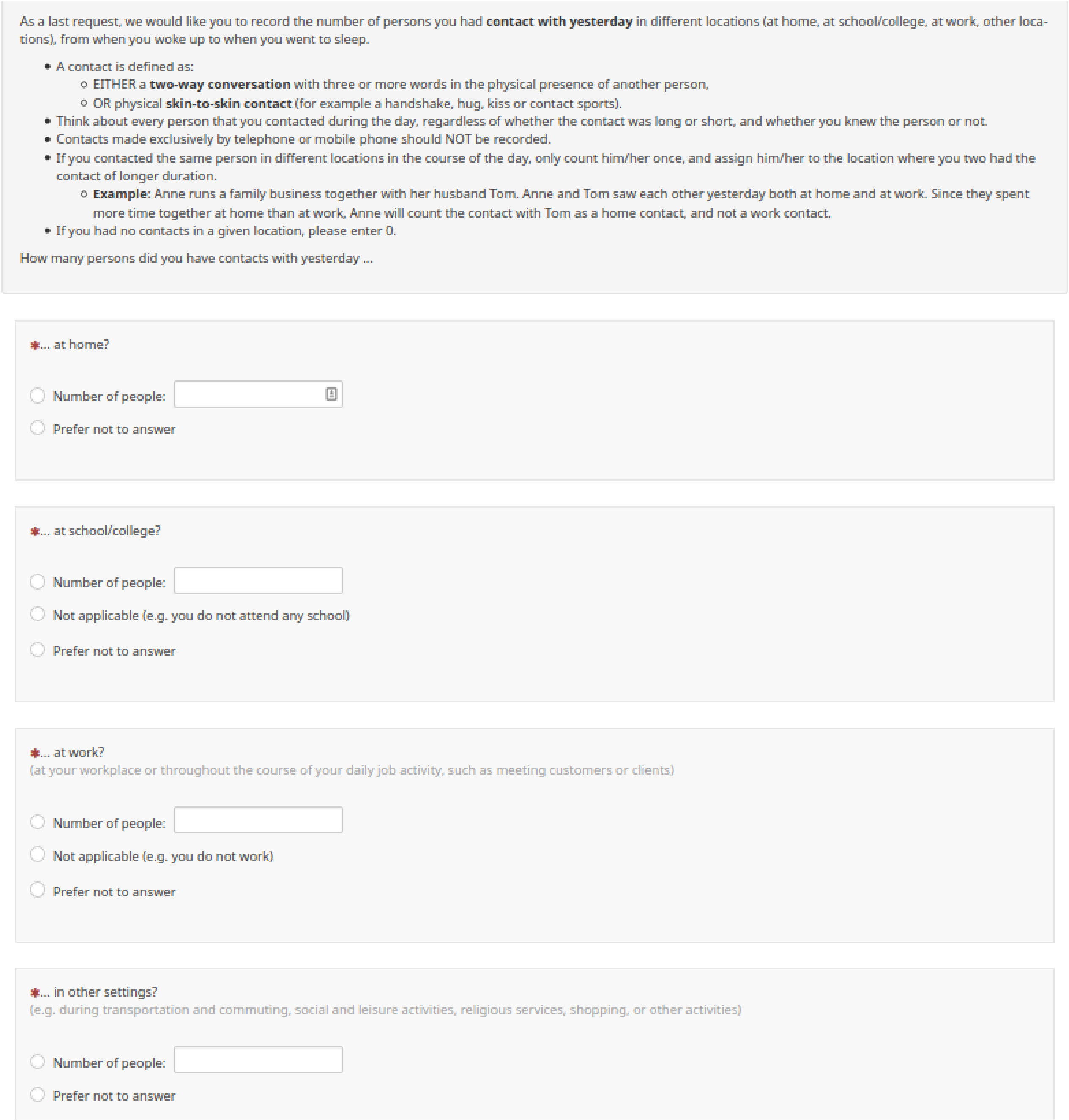

As a last request, we would like you to record the number of persons you had **contact with yesterday** in different locations (at home, at school/college, at work, other locations), from when you woke up to when you went to sleep.

- A contact is defined as:
  - EITHER a **two-way conversation** with three or more words in the physical presence of another person,
  - OR physical **skin-to-skin contact** (for example a handshake, hug, kiss or contact sports).
- Think about every person that you contacted during the day, regardless of whether the contact was long or short, and whether you knew the person or not.
- Contacts made exclusively by telephone or mobile phone should NOT be recorded.
- If you contacted the same person in different locations in the course of the day, only count him/her once, and assign him/her to the location where you two had the contact of longer duration.
  - **Example:** Anne runs a family business together with her husband Tom. Anne and Tom saw each other yesterday both at home and at work. Since they spent more time together at home than at work, Anne will count the contact with Tom as a home contact, and not a work contact.
- If you had no contacts in a given location, please enter 0.

How many persons did you have contacts with yesterday ...

*... at home?

- Number of people:
- Prefer not to answer

*... at school/college?

- Number of people:
- Not applicable (e.g. you do not attend any school)
- Prefer not to answer

*... at work?
(at your workplace or throughout the course of your daily job activity, such as meeting customers or clients)

- Number of people:
- Not applicable (e.g. you do not work)
- Prefer not to answer

*... in other settings?
(e.g. during transportation and commuting, social and leisure activities, religious services, shopping, or other activities)

- Number of people:
- Prefer not to answer

**Figure S1.** CHBS questions on daily social contacts.

### Additional results

#### Comparison between the raw and the weighted samples

In Tables S1-S8, for each country, we describe participants in terms of the variables used for the stratification and the construction of the Facebook advertising campaigns, comparing the unweighted sample to the weighted sample, and to the overall population.

We found that, in all countries, women were overrepresented in the sample Moreover, respondents were more uniformly spread across regions than in the real population. Finally, no trend in terms of age was found when comparing the raw and the adjusted sample distributions. We also noted that the unweighted sample size for the 65+ age group (who are at higher risk of death from COVID-19 [21], and who usually have lower social media participation rates [22]) was fairly large, with the Netherlands, UK, and the US showing higher percentages than in the population.

**Table S1.** Characteristics of survey participants in Belgium (N=3756), by region of residence, age group, and sex. We compare the raw sample ("unweighted") to the sample adjusted with the post-stratification weights ("weighted"), and to the distribution of each stratum in the population.

| **Variable** | **Category** | **Unweighted** | | **Weighted** | | **Population** |
|---|---|---|---|---|---|---|
| | | **N** | **%** | **N** | **%** | **%** |
| **Region** | **Brussels** | 685 | *18.2%* | 376 | *10.0%* | *10.3%* |
| | **Flanders** | 1552 | *41.3%* | 2220 | *59.2%* | *58.0%* |
| | **Wallonia** | 1519 | *40.4%* | 1157 | *30.8%* | *31.7%* |
| **Age group** | **18-24** | 673 | *17.9%* | 537 | *14.3%* | *13.7%* |
| | **25-44** | 1154 | *30.7%* | 1184 | *31.6%* | *31.2%* |
| | **45-64** | 1267 | *33.7%* | 1197 | *31.9%* | *32.4%* |
| | **65+** | 662 | *17.6%* | 835 | *22.2%* | *22.8%* |
| **Sex** | **Female** | 2579 | *68.7%* | 1948 | *51.9%* | *51.1%* |
| | **Male** | 1177 | *31.3%* | 1804 | *48.1%* | *48.9%* |

**Table S2.** Characteristics of survey participants in Germany (N=8584), by region of residence, age group, and sex. We compare the raw sample ("unweighted") to the sample adjusted with the post-stratification weights ("weighted"), and to the distribution of each stratum in the population.

| **Variable** | **Category** | **Unweighted** | | **Weighted** | | **Population** |
|---|---|---|---|---|---|---|
| | | **N** | **%** | **N** | **%** | **%** |
| **Region*** | **East** | 2095 | *24.4%* | 1477 | *17.2%* | *17.7%* |
| | **North** | 1985 | *23.1%* | 1555 | *18.1%* | *18.1%* |
| | **South** | 157 | *25.1%* | 2515 | *29.3%* | *29.0%* |
| | **West** | 2347 | *27.3%* | 3037 | *35.4%* | *35.2%* |
| **Age group** | **18-24** | 1866 | *21.7%* | 1019 | *11.9%* | *12.0%* |
| | **25-44** | 3323 | *38.7%* | 2513 | *29.3%* | *28.8%* |
| | **45-64** | 2249 | *26.2%* | 2909 | *33.9%* | *34.2%* |
| | **65+** | 1146 | *13.4%* | 2143 | *25.0%* | *24.9%* |
| **Sex** | **Female** | 5450 | *63.5%* | 4374 | *51.0%* | *51.0%* |
| | **Male** | 3134 | *36.5%* | 4210 | *49.0%* | *49.0%* |

* The "East" region includes Berlin, Brandenburg, Sachsen, Sachsen-Anhalt, and Thüringen; the "North" regions includes Bremen, Hamburg, Mecklenburg-Vorpommern, Niedersachsen, and Schleswig-Holstein; the "South" region includes Baden-Württemberg and Bayern; the "West" region includes Hessen, Nordrhein-Westfalen, Rheinland-Pfalz, and Saarland.

**Table S3.** Characteristics of survey participants in Spain (N=5360), by region of residence, age group, and sex. We compare the raw sample ("unweighted") to the sample adjusted with the post-stratification weights ("weighted"), and to the distribution of each stratum in the population.

| Variable | Category | Unweighted | | Weighted | | Population |
|---|---|---|---|---|---|---|
| | | N | % | N | % | % |
| **Region*** | **Central** | 656 | *12.2%* | 620 | *11.7%* | *11.9%* |
| | **East** | 1251 | *23.3%* | 1500 | *28.3%* | *29.1%* |
| | **Insular** | 417 | *7.8%* | 249 | *4.7%* | *4.8%* |
| | **Madrid** | 833 | *15.5%* | 755 | *14.3%* | *14.0%* |
| | **North East** | 620 | *11.6%* | 520 | *9.8%* | *9.6%* |
| | **North West** | 749 | *14.0%* | 507 | *9.6%* | *9.5%* |
| | **South** | 834 | *15.6%* | 1141 | *21.6%* | *21.1%* |
| **Age group** | **18-24** | 430 | *8.0%* | 606 | *11.5%* | *11.6%* |
| | **25-44** | 1994 | *37.2%* | 1676 | *31.7%* | *31.9%* |
| | **45-64** | 2175 | *40.6%* | 1821 | *34.4%* | *33.8%* |
| | **65+** | 761 | *14.2%* | 1188 | *22.5%* | *22.8%* |
| **Sex** | **Female** | 3764 | *70.2%* | 2769 | *52.3%* | *51.4%* |
| | **Male** | 1596 | *29.8%* | 2523 | *47.7%* | *48.6%* |

* The region "Central" includes Castilla y León, Castilla-la Mancha, and Extremadura; the region "East" includes Cataluña, Comunidad Valenciana, and Illes Balears; the region "Insular" includes Canarias; the region "North East" includes País Vasco, Comunidad Foral de Navarra, La Rioja, and Aragón; the region "North West" includes Galicia, Principado de Asturias, and Cantabria; the region "South" includes Andalucía, Región de Murcia, Ciudad Autónoma de Ceuta, and Ciudad Autónoma de Melilla.

**Table S4.** Characteristics of survey participants in France (N=4875), by region of residence, age group, and sex. We compare the raw sample ("unweighted") to the sample adjusted with the post-stratification weights ("weighted"), and to the distribution of each stratum in the population.

| Variable | Category | Unweighted | | Weighted | | Population |
|---|---|---|---|---|---|---|
| | | N | % | N | % | % |
| **Region*** | **Corse** | 17 | *0.4%* | 24 | *0.5%* | *0.5%* |
| | **Île de France** | 701 | *14.4%* | 906 | *18.7%* | *18.5%* |
| | **North East** | 1032 | *21.2%* | 1044 | *21.6%* | *22.0%* |
| | **South East** | 1010 | *20.7%* | 979 | *20.2%* | *20.2%* |
| | **South West** | 1043 | *21.4%* | 922 | *19.0%* | *18.7%* |
| | **West** | 1072 | *22.0%* | 968 | *20.0%* | *20.1%* |
| **Age group** | **18-24** | 876 | *18.0%* | 698 | *14.4%* | *14.2%* |
| | **25-44** | 1524 | *31.3%* | 1435 | *29.6%* | *29.4%* |
| | **45-64** | 1644 | *33.7%* | 1527 | *31.5%* | *31.6%* |
| | **65+** | 831 | *17.1%* | 1182 | *24.4%* | *24.7%* |
| **Sex** | **Female** | 3490 | *71.6%* | 2557 | *52.8%* | *52.2%* |
| | **Male** | 1385 | *28.4%* | 2285 | *47.2%* | *47.8%* |

* The region "North East" includes Bourgogne - Franche-Comté, Nord-Pas-de-Calais – Picardie, and Alsace - Champagne-Ardenne – Lorraine; the region "South East" includes Auvergne - Rhône-Alpes, and Provence-Alpes-Côte d'Azur; the region "South West" includes Aquitaine - Limousin - Poitou-Charentes, and Languedoc-Roussillon - Midi-Pyrénées; the region "West" includes Centre - Val de Loire, Normandie, Pays-de-la-Loire, and Bretagne.

**Table S5.** Characteristics of survey participants in Italy (N=7701), by region of residence, age group, and sex. We compare the raw sample ("unweighted") to the sample adjusted with the post-stratification weights ("weighted"), and to the distribution of each stratum in the population.

| Variable | Category | Unweighted | | Weighted | | Population |
|---|---|---|---|---|---|---|
| | | N | % | N | % | % |
| **Region*** | **Central** | 1640 | *21.3%* | 1521 | *19.9%* | *20.0%* |
| | **Insular** | 812 | *10.5%* | 810 | *10.6%* | *11.0%* |
| | **North East** | 1935 | *25.1%* | 1516 | *19.9%* | *19.3%* |
| | **North West** | 2311 | *30.0%* | 2025 | *26.5%* | *26.7%* |
| | **South** | 1003 | *13.0%* | 1761 | *23.1%* | *23.0%* |
| **Age group** | **18-24** | 1598 | *20.8%* | 834 | *10.9%* | *11.2%* |
| | **25-44** | 3152 | *40.9%* | 2187 | *28.7%* | *28.1%* |
| | **45-64** | 2072 | *26.9%* | 2609 | *34.2%* | *34.4%* |
| | **65+** | 879 | *11.4%* | 2003 | *26.2%* | *26.3%* |
| **Sex** | **Female** | 5042 | *65.5%* | 4030 | *52.8%* | *51.7%* |
| | **Male** | 2659 | *34.5%* | 3602 | *47.2%* | *48.3%* |

* The region "Central" includes Toscana, Umbria, Marche, and Lazio; the region "Insular" includes Sicilia and Sardegna; the region "North East" includes Provincia Autonoma di Bolzano/Bozen, Provincia Autonoma di Trento, Veneto, Friuli-Venezia Giulia, and Emilia-Romagna; the region "North West" includes Piemonte, Valle d'Aosta/Vallée d'Aoste, Liguria, and Lombardia; the region "South" includes Abruzzo, Molise, Campania, Puglia, Basilicata, and Calabria.

**Table S6.** Characteristics of survey participants in the Netherlands (N=3537), by region of residence, age group, and sex. We compare the raw sample ("unweighted") to the sample adjusted with the post-stratification weights ("weighted"), and to the distribution of each stratum in the population.

| Variable | Category | Unweighted | | Weighted | | Population |
|---|---|---|---|---|---|---|
| | | N | % | N | % | % |
| **Region*** | **East** | 879 | *24.9%* | 746 | *21.1%* | *20.9%* |
| | **North** | 735 | *20.8%* | 353 | *10.0%* | *10.1%* |
| | **South** | 888 | *25.1%* | 759 | *21.5%* | *21.5%* |
| | **West** | 1035 | *29.3%* | 1673 | *47.4%* | *47.5%* |
| **Age group** | **18-24** | 502 | *14.2%* | 537 | *15.2%* | *14.7%* |
| | **25-44** | 880 | *24.9%* | 1012 | *28.7%* | *29.3%* |
| | **45-64** | 1319 | *37.3%* | 1187 | *33.6%* | *33.3%* |
| | **65+** | 836 | *23.6%* | 795 | *22.5%* | *22.8%* |
| **Sex** | **Female** | 2370 | *67.0%* | 1802 | *51.0%* | *50.6%* |
| | **Male** | 1167 | *33.0%* | 1729 | *49.0%* | *49.4%* |

* The region "East" includes Overijssel, Gelderland, and Flevoland; the region "North" includes Groningen, Friesland, and Drenthe; the region "South" includes Noord-Brabant and Limburg; the region "West" includes Utrecht, Noord-Holland, Zuid-Holland, and Zeeland.

**Table S7.** Characteristics of survey participants in the United Kingdom (N=7416), by region of residence, age group, and sex. We compare the raw sample ("unweighted") to the sample adjusted with the post-stratification weights ("weighted"), and to the distribution of each stratum in the population.

| Variable | Category | Unweighted | | Weighted | | Population |
|---|---|---|---|---|---|---|
| | | N | % | N | % | % |
| **Region** | **England** | 3458 | *46.6%* | 5218 | *70.8%* | *70.9%* |
| | **London** | 774 | *10.4%* | 972 | *13.2%* | *13.2%* |
| | **Northern Ireland** | 545 | *7.4%* | 204 | *2.8%* | *2.8%* |
| | **Scotland** | 1506 | *20.3%* | 616 | *8.4%* | *8.4%* |
| | **Wales** | 1133 | *15.3%* | 356 | *4.8%* | *4.8%* |
| **Age group** | **18-24** | 595 | *8.0%* | 1018 | *13.8%* | *14.3%* |
| | **25-44** | 1846 | *24.9%* | 2355 | *32.0%* | *31.8%* |
| | **45-64** | 3070 | *41.4%* | 2332 | *31.7%* | *31.4%* |
| | **65+** | 1905 | *25.7%* | 1662 | *22.6%* | *22.4%* |
| **Sex** | **Female** | 4852 | *65.4%* | 3789 | *51.4%* | *51.0%* |
| | **Male** | 2564 | *34.6%* | 3578 | *48.6%* | *49.0%* |

**Table S8.** Characteristics of survey participants in the United States (N=12479), by region of residence, age group, and sex. We compare the raw sample ("unweighted") to the sample adjusted with the post-stratification weights ("weighted"), and to the distribution of each stratum in the population.

| **Variable** | **Category** | **Unweighted** | | **Weighted** | | **Population** |
|---|---|---|---|---|---|---|
| | | **N** | **%** | **N** | **%** | **%** |
| **Region*** | **Midwest** | 3359 | *26.9%* | 2571 | *20.7%* | *20.9%* |
| | **North East** | 2805 | *22.5%* | 2162 | *17.4%* | *17.5%* |
| | **South** | 3068 | *24.6%* | 4695 | *37.8%* | *37.8%* |
| | **West** | 3247 | *26.0%* | 2980 | *24.0%* | *23.8%* |
| **Age group** | **18-24** | 1387 | *11.1%* | 1923 | *15.5%* | *16.1%* |
| | **25-44** | 3394 | *27.2%* | 4059 | *32.7%* | *32.6%* |
| | **45-64** | 4254 | *34.1%* | 3950 | *31.8%* | *31.5%* |
| | **65+** | 3444 | *27.6%* | 2477 | *20.0%* | *19.7%* |
| **Sex** | **Female** | 8330 | *66.8%* | 6431 | *51.8%* | *51.2%* |
| | **Male** | 4149 | *33.3%* | 5977 | *48.2%* | *48.8%* |

* The region "Midwest" includes Illinois, Indiana, Iowa, Kansas, Michigan, Minnesota, Missouri, Nebraska, North Dakota, Ohio, South Dakota, and Wisconsin; the region "North East" includes Connecticut, Maine, Massachusetts, New Hampshire, New Jersey, New York, Pennsylvania, Rhode Island, and Vermont; the region "South" includes Alabama, Arkansas, Delaware, Florida, Georgia, Kentucky, Louisiana, Maryland, Mississippi, North Carolina, Oklahoma, South Carolina, Tennessee, Texas, Virginia, and West Virginia; the region "West" includes Alaska, Arizona, California, Colorado, Hawaii, Idaho, Montana, Nevada, New Mexico, Oregon, Utah, Washington, and Wyoming.

### Sensitivity analysis for the contact threshold values

Tables S9-S16 compare the contact threshold scenario presented in the manuscript, where individuals who reported more contacts than the 90% quantile of the contact distribution per setting were removed, with the scenario considered in the sensitivity analysis, where individuals who reported 30 or more contacts per setting were removed. For each scenario, we show the minimum, the median, and the first and the third quartile, and the maximum number of contacts for each setting and overall, as well as the number of missing values (either because the participant did not report it or because he or she reported a number of contacts above the considered threshold).

Under the 90% quantile scenario used in the main analysis, the number of missing values (due either to respondents who did not provide any contact number or who reported a number of contacts above the threshold) was much higher than under the $\leq 29$ scenario. Even though these thresholds might lead to different average contacts numbers, which is a metric not robust to outliers, the effect on the median and the quartiles was much smaller, with differences between the two outliers scenarios mainly found in Germany, the Netherlands, and the United States for the home contacts (and hence for the overall contacts).

**Table S9**. Contact distribution by threshold in each setting and overall, for Belgium (N=3756).

| Threshold | Statistic | Overall | Home | School | Work | General community |
|---|---|---|---|---|---|---|
| **90% quantile** | **Minimum** | 0 | 0 | 0 | 0 | 0 |
| | **First quartile** | 1 | 0 | 0 | 0 | 0 |
| | **Median** | 2 | 1 | 0 | 0 | 0 |
| | **Third quartile** | 4 | 2 | 0 | 0 | 1 |
| | **Maximum** | 28 | 4 | 0 | 25 | 5 |
| | **No. missing values** | 530 | 185 | 50 | 142 | 212 |
| $\leq 29$ | **Minimum** | 0 | 0 | 0 | 0 | 0 |
| | **First quartile** | 1 | 0 | 0 | 0 | 0 |
| | **Median** | 2 | 1 | 0 | 0 | 0 |
| | **Third quartile** | 4 | 2 | 0 | 0 | 1 |
| | **Maximum** | 45 | 13 | 17 | 26 | 29 |
| | **No. missing values** | 156 | 2 | 2 | 139 | 23 |

**Table S10.** Contact distribution by threshold in each setting and overall, for Germany (N=8584).

| Threshold | Statistic | Overall | Home | School | Work | General community |
|---|---|---|---|---|---|---|
| **90% quantile** | **Minimum** | 0 | 0 | 0 | 0 | 0 |
| | **First quartile** | 1 | 1 | 0 | 0 | 0 |
| | **Median** | 3 | 1 | 0 | 0 | 0 |
| | **Third quartile** | 5 | 2 | 0 | 1 | 2 |
| | **Maximum** | 34 | 4 | 0 | 25 | 5 |
| | **No. Missing values** | 1653 | 530 | 98 | 424 | 904 |
| $\leq 29$ | **Minimum** | 0 | 0 | 0 | 0 | 0 |
| | **First quartile** | 2 | 1 | 0 | 0 | 0 |
| | **Median** | 4 | 1 | 0 | 0 | 1 |
| | **Third quartile** | 7 | 3 | 0 | 1 | 2 |
| | **Maximum** | 61 | 29 | 24 | 28 | 27 |
| | **No. Missing values** | 540 | 14 | 8 | 416 | 170 |

**Table S11.** Contact distribution by threshold in each setting and overall, for Spain (N=5360).

| Threshold | Statistic | Overall | Home | School | Work | General community |
|---|---|---|---|---|---|---|
| **90% quantile** | **Minimum** | 0 | 0 | 0 | 0 | 0 |
| | **First quartile** | 1 | 1 | 0 | 0 | 0 |
| | **Median** | 2 | 1 | 0 | 0 | 0 |
| | **Third quartile** | 3 | 3 | 0 | 0 | 0 |
| | **Maximum** | 31 | 4 | 0 | 25 | 5 |
| | **No. Missing values** | 609 | 213 | 82 | 148 | 244 |
| $\leq 29$ | **Minimum** | 0 | 0 | 0 | 0 | 0 |
| | **First quartile** | 1 | 1 | 0 | 0 | 0 |
| | **Median** | 2 | 2 | 0 | 0 | 0 |
| | **Third quartile** | 4 | 3 | 0 | 0 | 1 |
| | **Maximum** | 76 | 28 | 25 | 28 | 25 |
| | **No. Missing values** | 198 | 12 | 7 | 145 | 56 |

**Table S12.** Contact distribution by threshold in each setting and overall, for France (N=4875).

| Threshold | Statistic | Overall | Home | School | Work | General community |
|---|---|---|---|---|---|---|
| **90% quantile** | **Minimum** | 0 | 0 | 0 | 0 | 0 |
| | **First quartile** | 1 | 0 | 0 | 0 | 0 |
| | **Median** | 2 | 1 | 0 | 0 | 0 |
| | **Third quartile** | 3 | 2 | 0 | 0 | 0 |
| | **Maximum** | 32 | 4 | 0 | 25 | 5 |
| | **No. Missing values** | 591 | 166 | 66 | 179 | 258 |
| **≤ 29** | **Minimum** | 0 | 0 | 0 | 0 | 0 |
| | **First quartile** | 1 | 0 | 0 | 0 | 0 |
| | **Median** | 2 | 1 | 0 | 0 | 0 |
| | **Third quartile** | 4 | 2 | 0 | 0 | 1 |
| | **Maximum** | 54 | 18 | 25 | 28 | 25 |
| | **No. Missing values** | 223 | 4 | 7 | 176 | 62 |

**Table S13.** Contact distribution by threshold in each setting and overall, for Italy (N=7701).

| Threshold | Statistic | Overall | Home | School | Work | General community |
|---|---|---|---|---|---|---|
| **90% quantile** | **Minimum** | 0 | 0 | 0 | 0 | 0 |
| | **First quartile** | 1 | 1 | 0 | 0 | 0 |
| | **Median** | 2 | 2 | 0 | 0 | 0 |
| | **Third quartile** | 4 | 3 | 0 | 0 | 0 |
| | **Maximum** | 29 | 4 | 0 | 25 | 5 |
| | **No. Missing values** | 781 | 387 | 83 | 169 | 220 |
| **≤ 29** | **Minimum** | 0 | 0 | 0 | 0 | 0 |
| | **First quartile** | 1 | 1 | 0 | 0 | 0 |
| | **Median** | 2 | 2 | 0 | 0 | 0 |
| | **Third quartile** | 4 | 3 | 0 | 0 | 1 |
| | **Maximum** | 42 | 22 | 22 | 25 | 23 |
| | **No. Missing values** | 195 | 12 | 5 | 169 | 24 |

**Table S14.** Contact distribution by threshold in each setting and overall, for the Netherlands (N=3537).

| Threshold | Statistic | Overall | Home | School | Work | General community |
|---|---|---|---|---|---|---|
| **90% quantile** | **Minimum** | 0 | 0 | 0 | 0 | 0 |
| | **First quartile** | 1 | 1 | 0 | 0 | 0 |
| | **Median** | 3 | 1 | 0 | 0 | 0 |
| | **Third quartile** | 5 | 2 | 0 | 0 | 2 |
| | **Maximum** | 33 | 4 | 0 | 25 | 5 |
| | **No. Missing values** | 569 | 237 | 50 | 124 | 249 |
| $\leq 29$ | **Minimum** | 0 | 0 | 0 | 0 | 0 |
| | **First quartile** | 1 | 1 | 0 | 0 | 0 |
| | **Median** | 3 | 1 | 0 | 0 | 0 |
| | **Third quartile** | 7 | 3 | 0 | 0 | 2 |
| | **Maximum** | 46 | 20 | 18 | 29 | 25 |
| | **No. Missing values** | 151 | 9 | 4 | 120 | 29 |

**Table S15.** Contact distribution by threshold in each setting and overall, for the UK (N=7416).

| Threshold | Statistic | Overall | Home | School | Work | General community |
|---|---|---|---|---|---|---|
| **90% quantile** | **Minimum** | 0 | 0 | 0 | 0 | 0 |
| | **First quartile** | 1 | 1 | 0 | 0 | 0 |
| | **Median** | 2 | 1 | 0 | 0 | 0 |
| | **Third quartile** | 4 | 2 | 0 | 0 | 1 |
| | **Maximum** | 34 | 4 | 0 | 25 | 5 |
| | **No. Missing values** | 1357 | 437 | 178 | 361 | 711 |
| $\leq 29$ | **Minimum** | 0 | 0 | 0 | 0 | 0 |
| | **First quartile** | 1 | 1 | 0 | 0 | 0 |
| | **Median** | 2 | 1 | 0 | 0 | 0 |
| | **Third quartile** | 5 | 3 | 0 | 0 | 2 |
| | **Maximum** | 79 | 25 | 25 | 29 | 29 |
| | **No. Missing values** | 479 | 16 | 35 | 353 | 170 |

**Table S16.** Contact distribution by threshold in each setting and overall, for the US (N=12,479).

| Threshold | Statistic | Overall | Home | School | Work | General community |
|---|---|---|---|---|---|---|
| **90% quantile** | **Minimum** | 0 | 0 | 0 | 0 | 0 |
| | **First quartile** | 1 | 1 | 0 | 0 | 0 |
| | **Median** | 2 | 1 | 0 | 0 | 0 |
| | **Third quartile** | 4 | 2 | 0 | 0 | 1 |
| | **Maximum** | 32 | 4 | 0 | 25 | 5 |
| | **No. Missing values** | 2698 | 1098 | 204 | 589 | 1377 |
| $\leq 29$ | **Minimum** | 0 | 0 | 0 | 0 | 0 |
| | **First quartile** | 1 | 1 | 0 | 0 | 0 |
| | **Median** | 3 | 1 | 0 | 0 | 0 |
| | **Third quartile** | 6 | 3 | 0 | 0 | 2 |
| | **Maximum** | 60 | 28 | 25 | 29 | 28 |
| | **No. Missing values** | 851 | 28 | 48 | 585 | 316 |

#### Distribution of contact numbers

Two features of the highly skewed distribution of the overall social contacts (Figure S2) are worth noting. First, we found that the percentages of participants who reported having fewer than one contact per day were relatively high, at more than 35% in all countries except for Germany, the Netherlands, and the US. Second, the distribution was characterized by a long right tail, the length of which depended on the threshold used to remove the outliers at the top of the distribution (Tables S9-S16).

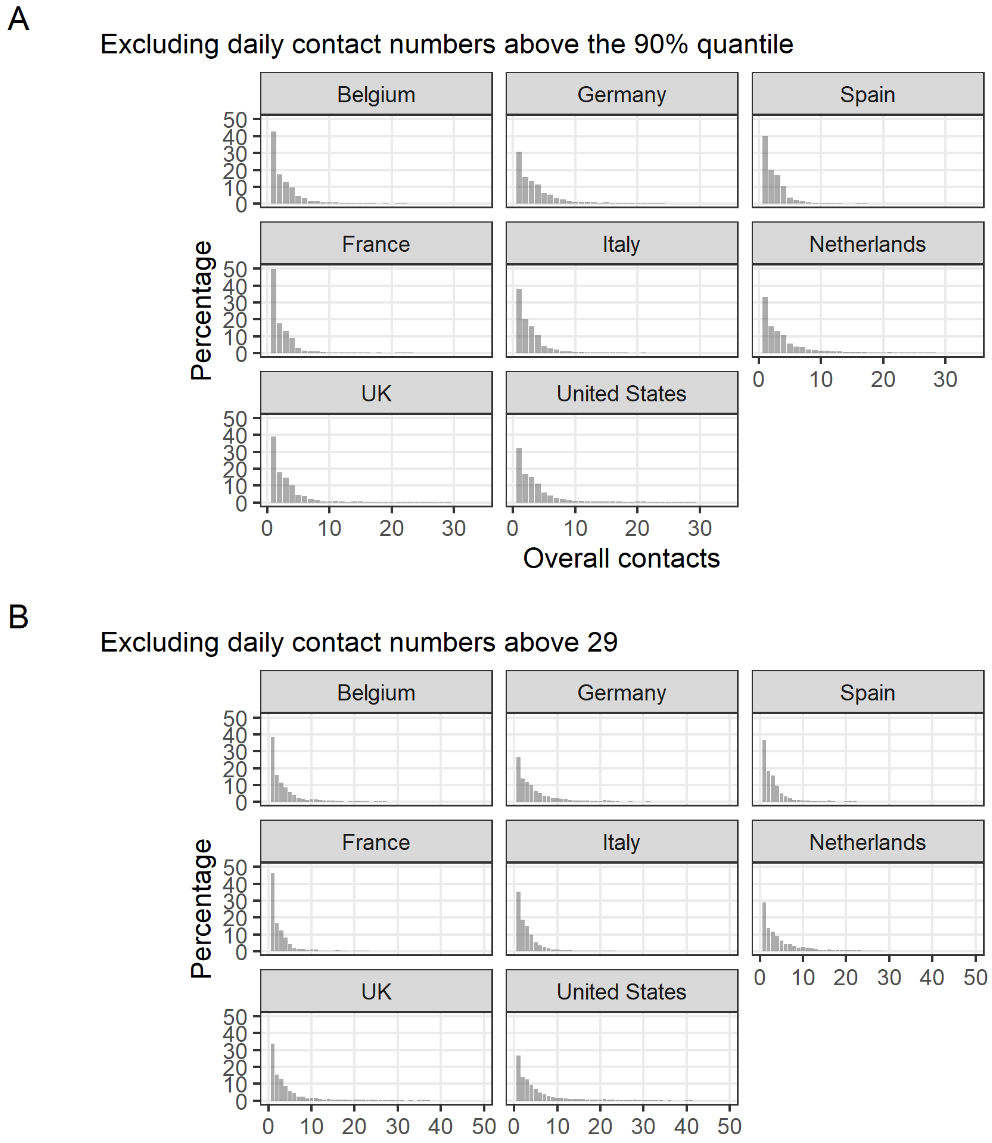


**Figure S2.** Histogram with the weighted distribution of the overall number of contacts by country. All contacts exclude the top 10% (A) and those equal to 30 or more (B).

### Contact numbers by setting and week

In Figures S3-S4, we showed the evolution of the weekly average number of contacts by setting, after applying the post-stratification weights, under the two threshold scenarios. As expected, home and school contact numbers showed little to no variability, with the former remaining almost constant during the whole study period, and the latter being reduced to zero due to school closures in every surveyed country.

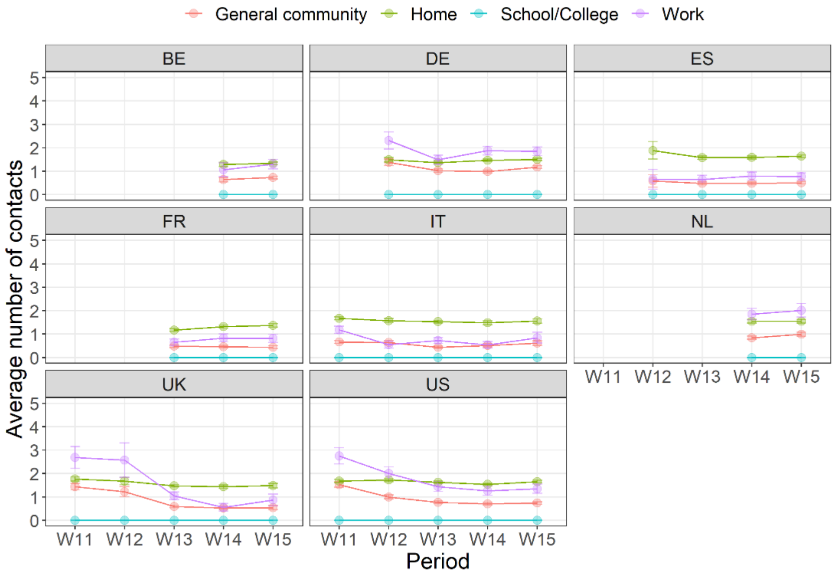


**Figure S3.** Average number of daily social contacts by setting, week, and country. Respondents with contacts above the 90% quantile threshold were removed from the analysis.

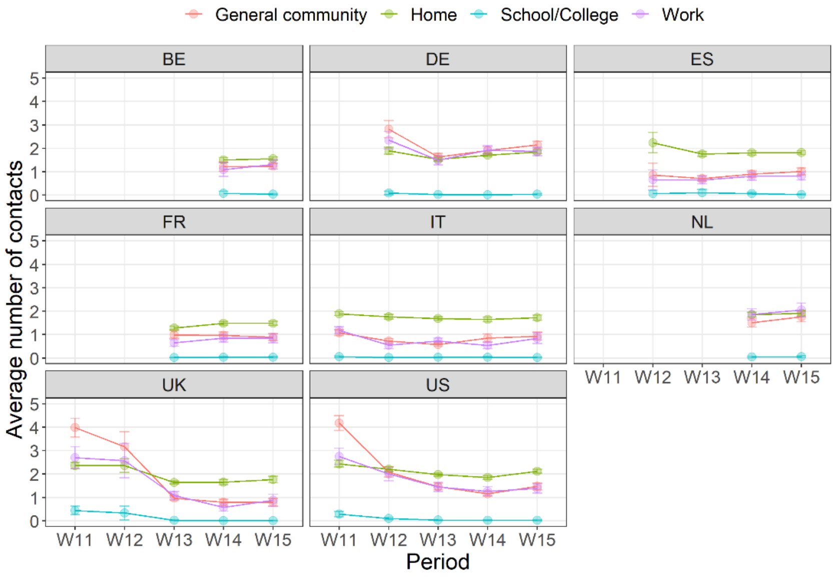


**Figure S4.** Average number of daily social contacts by setting, week, and country. Respondents with contacts above the $\leq \mathbf{29}$ threshold were removed from the analysis.

### Household size and home contacts

Figure S5 shows the consistency of the household size and the number of home contacts under the two thresholds. More than half of respondents lived either alone or with another person. Respondents living in large households, with five members or more, were more frequent in the US (around 20%). Home contacts ranged between around half a contact per day, reported by people living alone, to three contacts per day or more, reported by people living in households of four or more components.

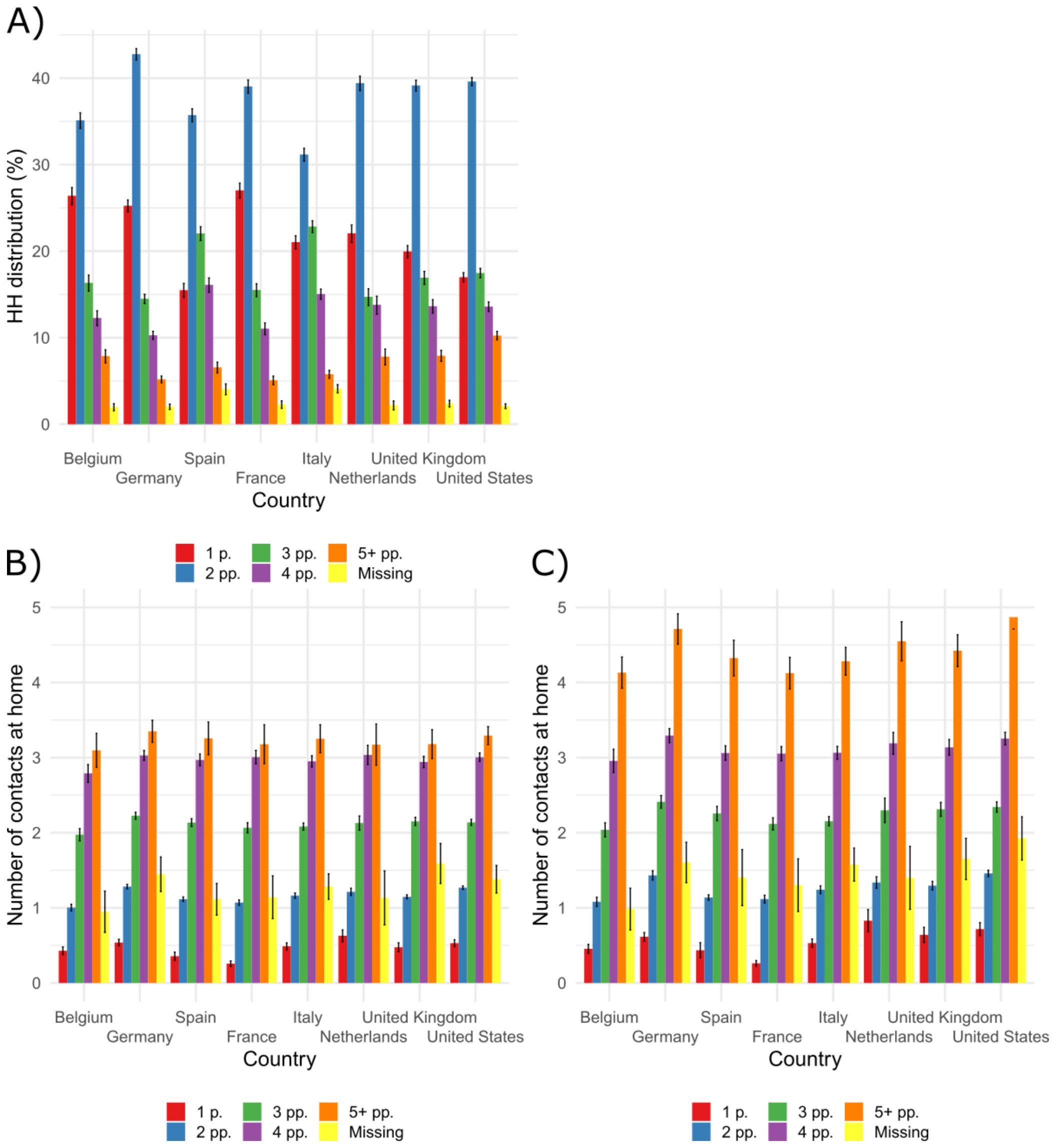


**Figure S5.** Household size distribution by country (A), compared to the average number of contacts at home by household size and country, under the 90% quantile threshold (B), and under the $\leq \mathbf{29}$ threshold (C).

### Contact numbers by age group and week at work and in the general community

Figures S6 and S7 show in panel A the weekly evolution of the predicted number of contacts at work and in the general community, respectively, in the pre-COVID period [10, 11] and during our study, stratified by age group. In panel B, we show the comparison between the values in the pre-COVID period and those in the first week available for each country. The United States and Spain are excluded from panel B, as no pre-COVID data are available for these two countries.

When we looked at the trends in work contact numbers (Figure S6), we found that the 65+ age group reported having fewer contacts than the other age groups, except in Belgium and France. When we focused on the countries with the longest observation time periods, we noticed that among the youngest age groups in the UK, there was an overall decrease in work contact numbers throughout the study period that might reflect the implementation of physical distancing measures over time. By contrast, when we examined countries such as France, Spain and Italy, we found that the younger age groups already had few to no contacts, which is likely because in these countries, the full lockdown measures were already preventing a large portion of the population from going to work.

The trends for contact numbers in the general community did not differ significantly by age group, or by week (Figure S7). Moreover, these contact numbers had already reached their lowest levels by the beginning of the study period in each country, except in the UK and the US. This evidence might suggest that, except in the latter two countries, the big drop in the overall contacts shown in Fig 3 in the manuscript is mostly attributable to a reduction in work contact numbers, and not to a reduction in contact numbers in the general community.

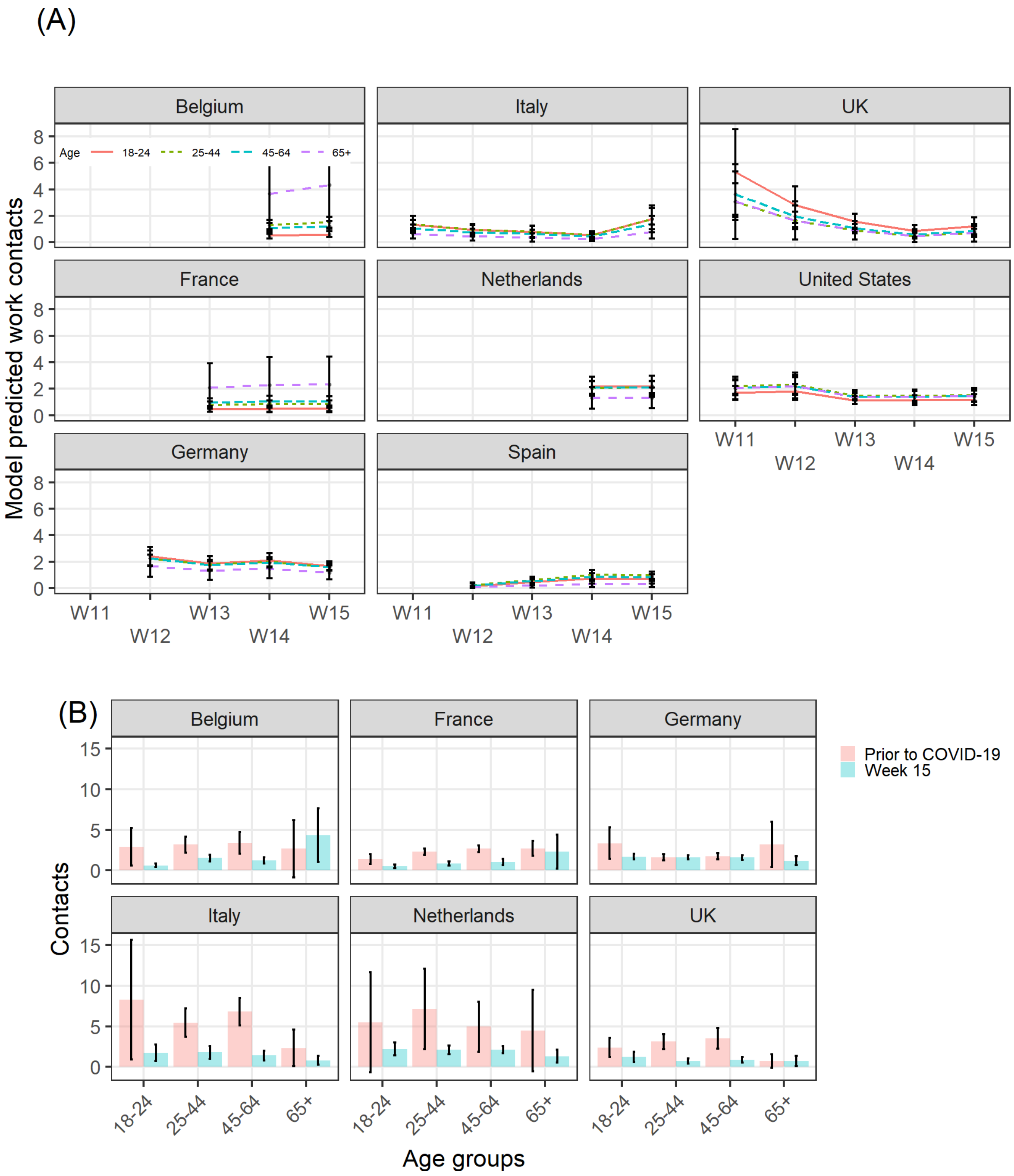


**Figure S6.** Model-predicted daily number of social contacts at work, by age group, country, and week, March-April 2020. (A). Comparison of model-predicted overall contact numbers between the pre-COVID period and calendar week 15, by country (B). Respondents with contacts above the 90% quantile threshold were removed from the analysis.

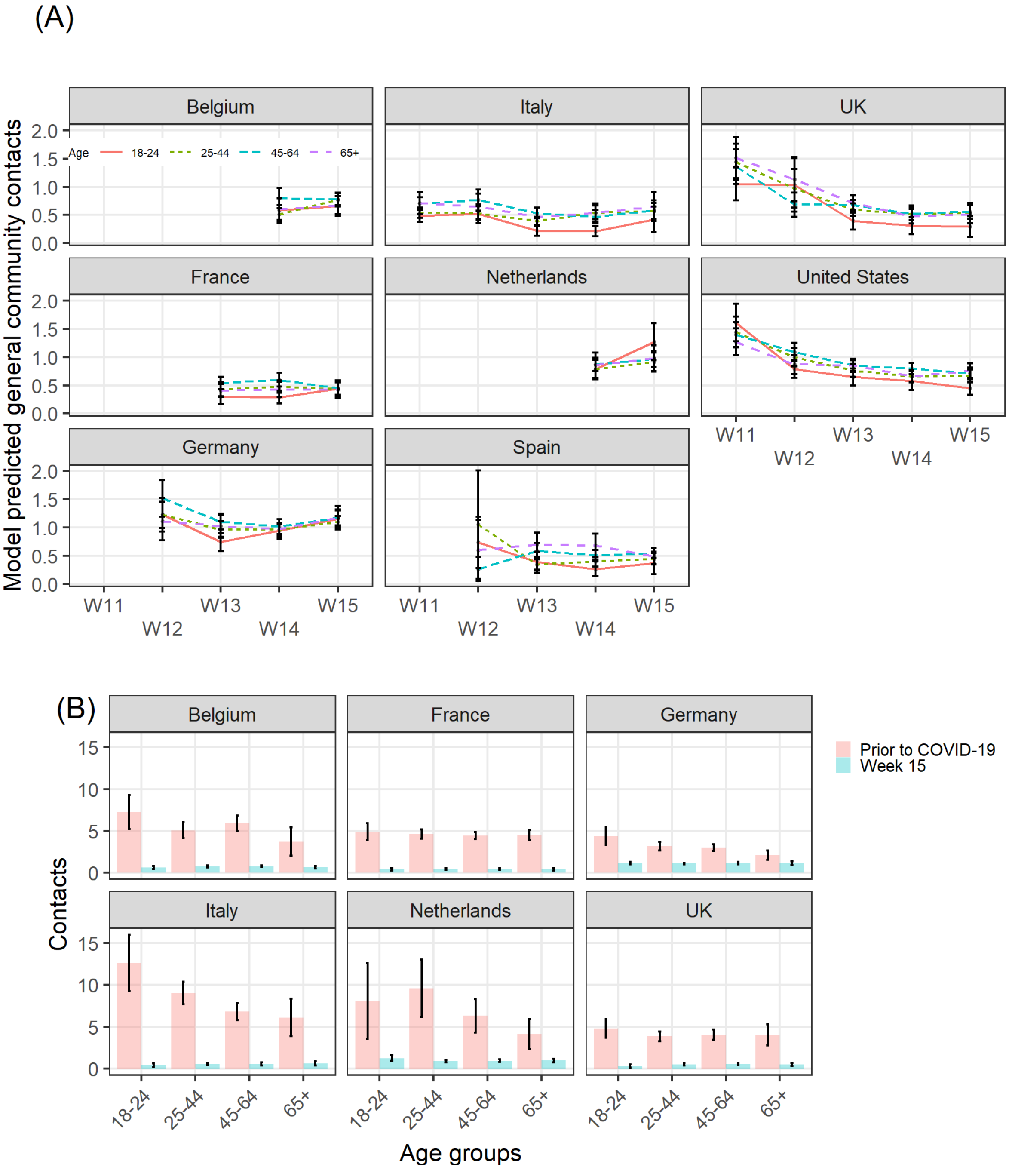


**Figure S7.** Model-predicted daily number of social contacts in the general community, by age group, country, and week, March-April 2020. (A). Comparison of model-predicted overall contact numbers between the pre-COVID period and calendar week 15, by country (B). Respondents with contacts above the 90% quantile threshold were removed from the analysis.

### Sensitivity analysis

We performed a sensitivity analysis of the threshold chosen to curtail the right tail of the distribution of social contacts, in response to the extremely high values reported by some of the participants. In the main analysis, we presented the results of the threshold corresponding to the 90% quantile of the distribution of the setting-specific contacts (removing the respondents with more contacts than the threshold from the analysis sample), which were subsequently summed up to provide the overall count of contacts. In the following figures and tables, we present the results of the analysis using a fixed cut-off point at 29 contacts per day, removing respondents reporting 30 contacts per day or more in each of the settings.

Figure S8 shows the daily pattern of contacts, overall and in different settings (home, work, and general community). The time patterns per country are like those shown by the 90% quantile threshold, shown in Fig 1 in the manuscript, even though the daily numbers and the weekly estimates (Table S17) are generally higher by about one contact per day per setting.

Figures S9 to S11 show the estimates of the daily contact numbers by week, overall, at work, and in the general community, disaggregated by age group. We found that contact numbers in the last study week (week 15, April 6 – 12) were indeed lower than those reported by the same age groups in the period prior to COVID-19 in almost all countries, even though not as much as with the 90% quantile threshold. The exception was Germany, as for most of the age groups, especially the older adults, we did not find evidence of a reduction in social contacts compared to the pre-pandemic period, regardless of the setting.

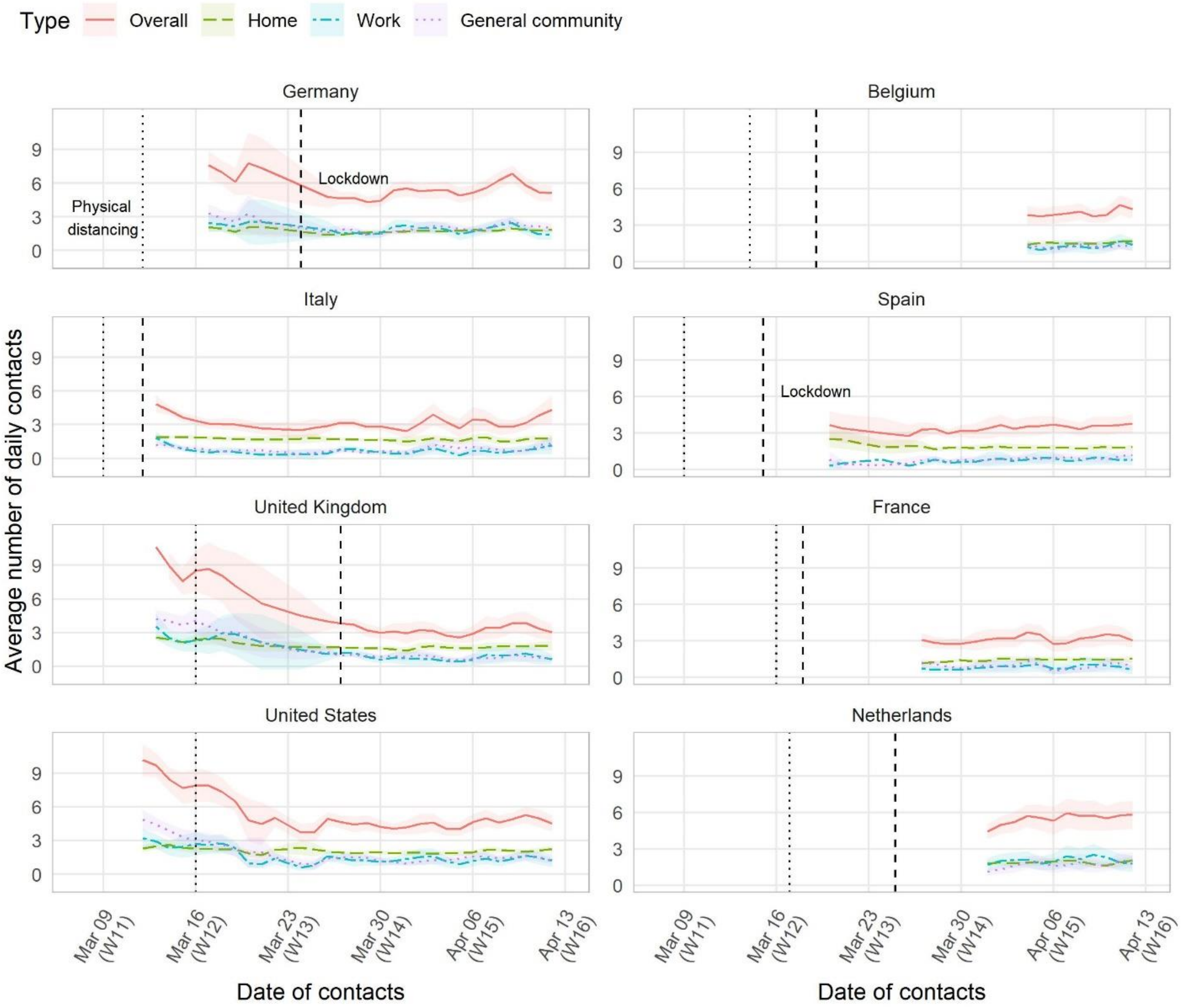


**Figure S8.** Mean overall number of daily social contacts (with 95% CI), smoothed by a simple two-day moving average, by country and study day. The dotted line corresponds to the date in which the physical distancing guidelines were introduced at the national level; the dashed line corresponds to the date in which the lockdown, regardless of being full or partial, was ordered. Respondents with contacts above the $\leq \mathbf{29}$ threshold were removed from the analysis.

**Table S17.** Model-predicted mean number (with SE) of daily contacts per person compared with pre-pandemic model predictions, by country, setting, and week, March-April 2020. Respondents with contacts above the $\leq \mathbf{29}$ threshold were removed from the analysis.

| Country | Setting | Prior to COVID-19 | | Week 11 (Mar 9 – 15) | | Week 12 (Mar 16 – 22) | | Week 13 (March 23 – 29) | | Week 14 (Mar 30 - Apr 5) | | Week 15 (Apr 6 – 12) | |
|---|---|---|---|---|---|---|---|---|---|---|---|---|---|
| | | Mean | SE | Mean | SE | Mean | SE | Mean | SE | Mean | SE | Mean | SE |
| Belgium | Work | 3.2 | 0.41 | | | | | | | 1.15 | 0.15 | 1.34 | 0.11 |
| | General community | 5.33 | 0.29 | | | | | | | 1.21 | 0.11 | 1.24 | 0.06 |
| | Overall | 11.76 | 0.48 | | | | | | | 3.82 | 0.22 | 4.06 | 0.14 |
| France | Work | 2.37 | 0.12 | | | | | 0.71 | 0.1 | 0.87 | 0.1 | 0.85 | 0.1 |
| | General community | 4.59 | 0.13 | | | | | 0.91 | 0.08 | 1.06 | 0.09 | 0.89 | 0.07 |
| | Overall | 10.3 | 0.21 | | | | | 2.97 | 0.14 | 3.2 | 0.14 | 3.09 | 0.13 |
| Germany | Work | 1.83 | 0.17 | | | 2.15 | 0.18 | 1.77 | 0.13 | 1.81 | 0.09 | 1.78 | 0.09 |
| | General community | 2.98 | 0.14 | | | 2.8 | 0.19 | 1.63 | 0.08 | 1.91 | 0.08 | 2.14 | 0.08 |
| | Overall | 8.03 | 0.27 | | | 6.47 | 0.29 | 4.84 | 0.16 | 5.21 | 0.13 | 5.59 | 0.14 |
| Italy | Work | 5.72 | 0.62 | 1.16 | 0.1 | 0.88 | 0.17 | 0.68 | 0.07 | 0.51 | 0.07 | 1.13 | 0.19 |
| | General community | 7.93 | 0.36 | 0.96 | 0.06 | 0.85 | 0.08 | 0.58 | 0.04 | 0.89 | 0.09 | 1.05 | 0.09 |
| | Overall | 18.22 | 0.61 | 3.97 | 0.12 | 3.33 | 0.15 | 2.9 | 0.09 | 3.17 | 0.17 | 3.82 | 0.2 |
| Netherlands | Work | 5.83 | 1.31 | | | | | | | 1.88 | 0.14 | 2.23 | 0.19 |
| | General community | 7.13 | 0.76 | | | | | | | 1.49 | 0.09 | 1.77 | 0.11 |
| | Overall | 15.03 | 1.2 | | | | | | | 4.99 | 0.17 | 5.65 | 0.21 |
| Spain | Work | | | | | 0.75 | 0.25 | 0.63 | 0.09 | 0.89 | 0.1 | 0.84 | 0.09 |
| | General community | | | | | 0.84 | 0.22 | 0.73 | 0.06 | 0.88 | 0.07 | 1.03 | 0.08 |
| | Overall | | | | | 3.22 | 0.37 | 3.2 | 0.14 | 3.46 | 0.15 | 3.62 | 0.14 |
| United Kingdom | Work | 3.02 | 0.39 | 3.06 | 0.31 | 2.53 | 0.5 | 1.07 | 0.1 | 0.66 | 0.11 | 0.96 | 0.17 |
| | General community | 4.06 | 0.21 | 4.57 | 0.28 | 2.69 | 0.32 | 1.01 | 0.06 | 0.71 | 0.06 | 0.76 | 0.08 |
| | Overall | 10.54 | 0.35 | 9.71 | 0.41 | 7.46 | 0.64 | 3.61 | 0.12 | 2.93 | 0.14 | 3.37 | 0.17 |
| United States | Work | | | 2.86 | 0.2 | 1.93 | 0.14 | 1.43 | 0.1 | 1.23 | 0.1 | 1.38 | 0.11 |
| | General community | | | 4.2 | 0.17 | 2.12 | 0.11 | 1.45 | 0.07 | 1.16 | 0.06 | 1.48 | 0.07 |
| | Overall | | | 9.24 | 0.28 | 6.17 | 0.2 | 4.69 | 0.13 | 4.27 | 0.13 | 4.97 | 0.14 |

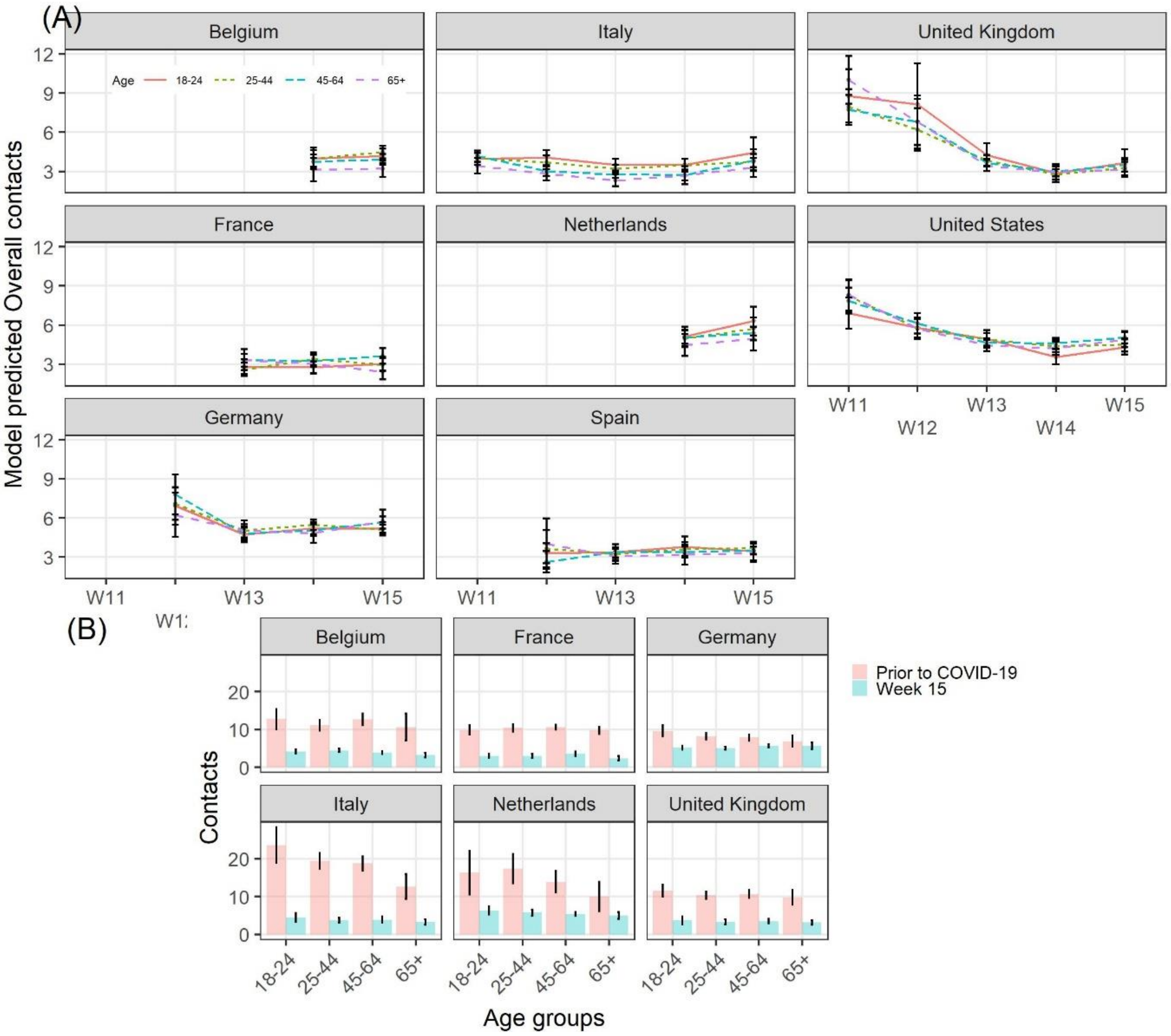


**Figure S9** .Model-predicted overall daily number of social contacts, by age group, country, and week, March-April 2020. (A). Comparison of model-predicted overall contact numbers between the pre-COVID period and calendar week 15, by country (B). Respondents with contacts above the $\leq \mathbf{29}$ threshold were removed from the analysis.

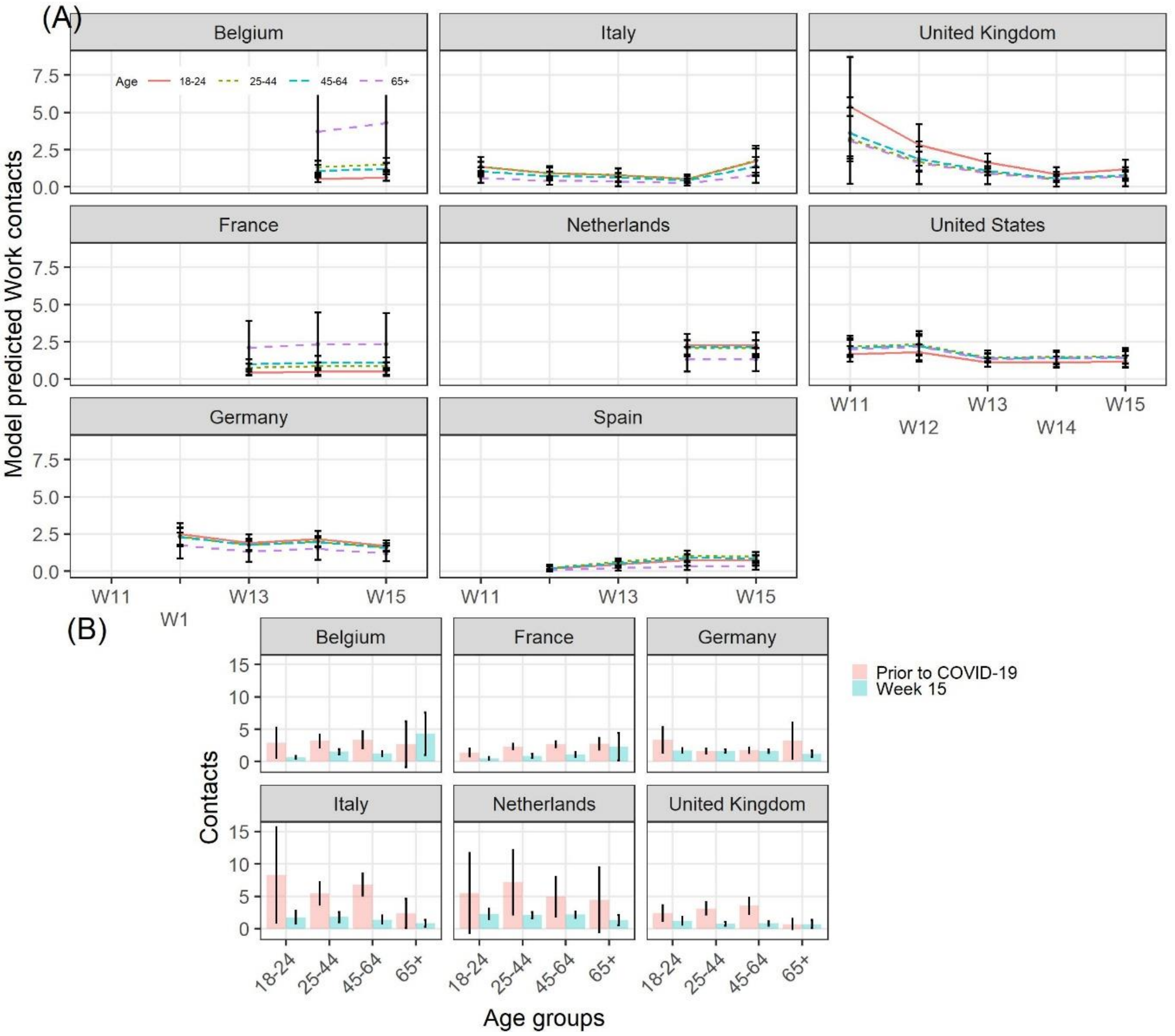


**Figure S10.** Model-predicted daily number of social contacts at work, by age group, country, and week, March-April 2020. (A). Comparison of model-predicted overall contact numbers between the pre-COVID period and calendar week 15, by country (B). Respondents with contacts above the $\leq \mathbf{29}$ threshold were removed from the analysis.

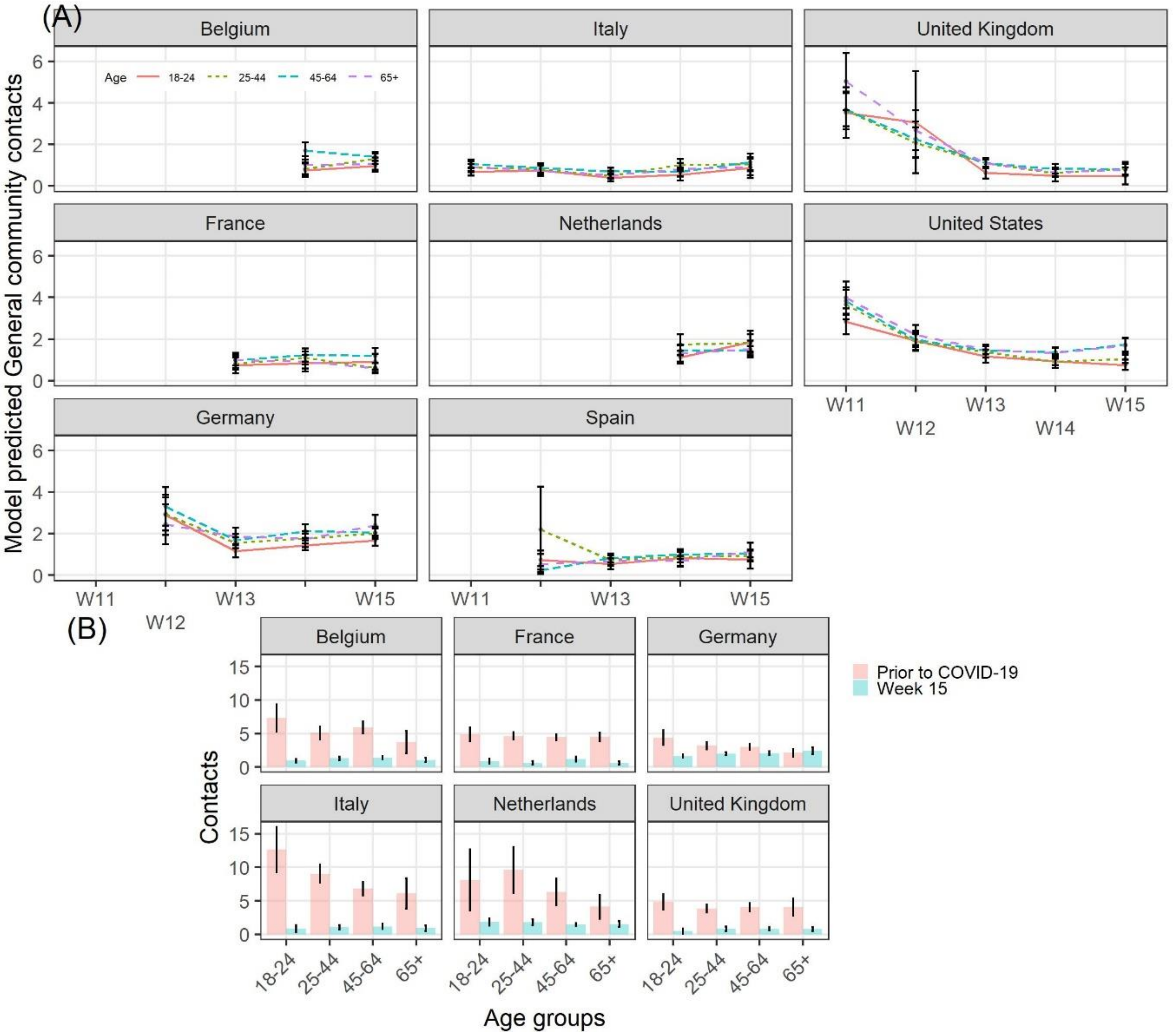


**Figure S11.** Model-predicted daily number of social contacts in the general community, by age group, country, and week, March-April 2020. (A). Comparison of model-predicted overall contact numbers between the pre-COVID period and calendar week 15, by country (B). Respondents with contacts above the $\leq \mathbf{29}$ threshold were removed from the analysis.

Finally, Figure S12 reports on the impact of the reduction of the age-specific social contacts on age mixing and hence on the net reproduction number $R_t$ on the different study weeks. Results are consistent with those for the 90% quantile threshold, as we found evidence of a reduction of the net reproduction number across the weeks, even though not as much as with the other threshols. The 95% CIs around the values of the $R_t$ in Italy, Spain, and in the UK (on week 14) were entirely below zero, as we observed with the 90% quantile threshold too, while in Belgium and France the 95% CI included one (as well as in the Netherlands and the US). For Germany, the $R_t$ was always above one, as a consequence of the small reduction in social contacts compared to the period prior to COVID-19.

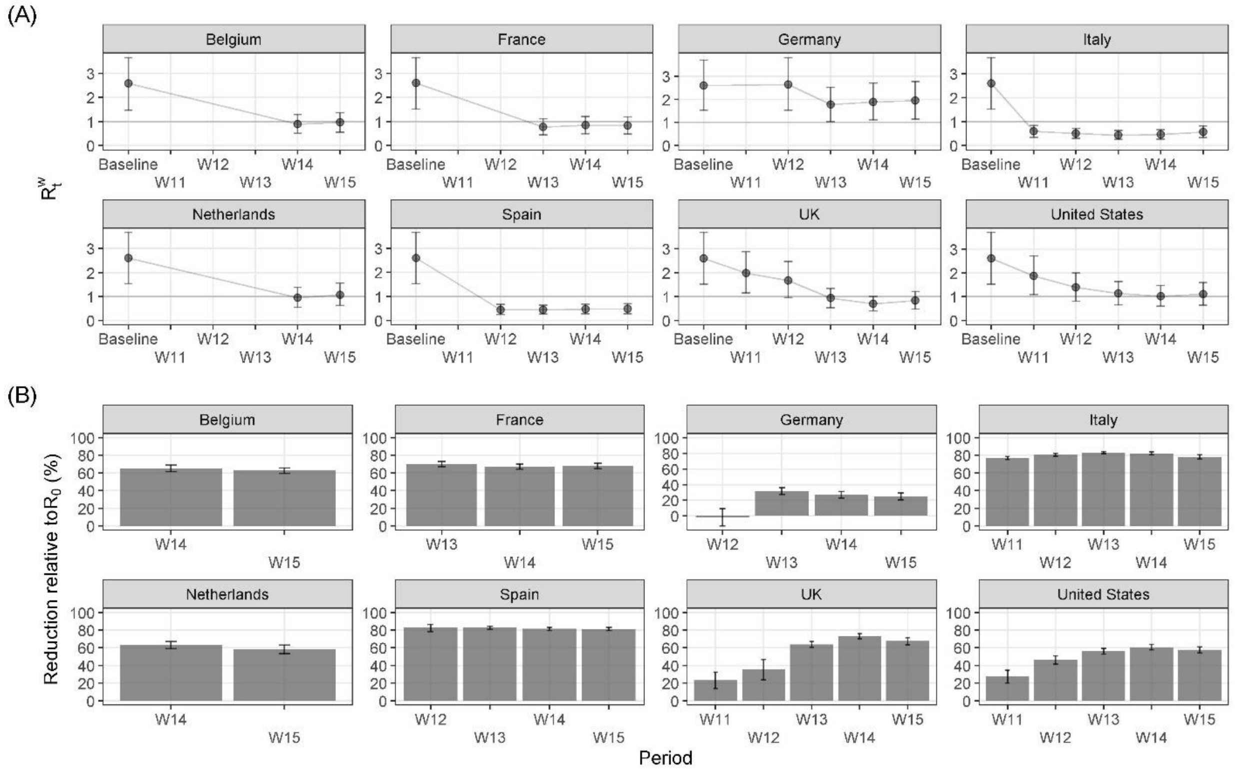


**Figure S12.** Absolute change in the weekly net reproduction number $R_t^w$ with respect to the basic reproduction number $R_0$ at baseline (A) and percent reduction with respect to R_0 (B), by country and week. The 95% CIs are based on 5,000 replicates. Respondents with contacts above the $\leq \mathbf{29}$ threshold were removed from the analysis.

### Association between behavioral change and social contact numbers

Figures S13 – S16 show the exponentiated coefficients of the health behaviors and the feeling of threat from the negative binomial regression model to estimate the overall number of daily contacts by study week, under the two different contact thresholds. For each country and variable, we show the main effect for the association between each variable and the overall number of social contacts, as well as how this association changed across the study weeks.

Two results are worthy to highlight. First, we found some evidence that people living in countries where the use of face masks increased over time reported higher number of social contacts (Figure S14). Such a result was reported for the US (from the end of March till the end of the study period), Italy (at least under the 90% quantile threshold), and Belgium. Second, we found a negative effect of the avoidance of social activities on social contact numbers in the Netherlands, the UK, and the US, where at the beginning only milder non-pharmaceutical interventions were implemented, with no obligation of avoiding social activities (Figure S15).

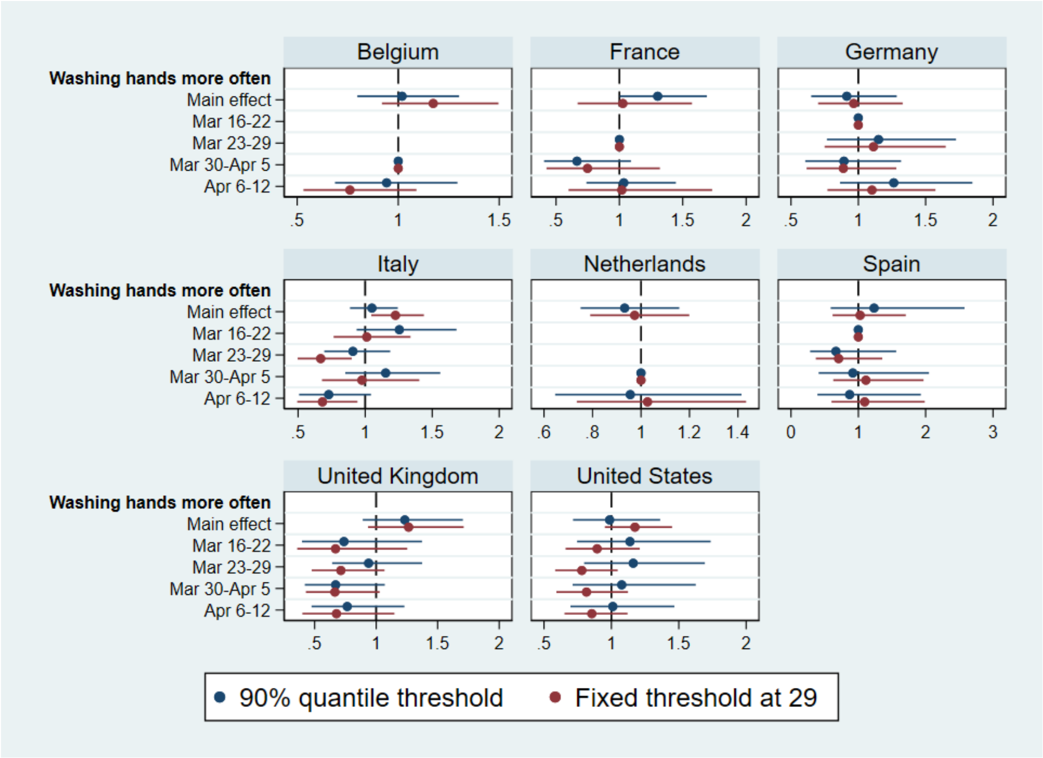


**Figure S13.** Effect of washing hand more often on overall contacts by study week. Exponentiated coefficients for the protective action "Washing hands more often", measuring the association between the action and the overall number of social contacts across study weeks. Results for both contact thresholds are shown.

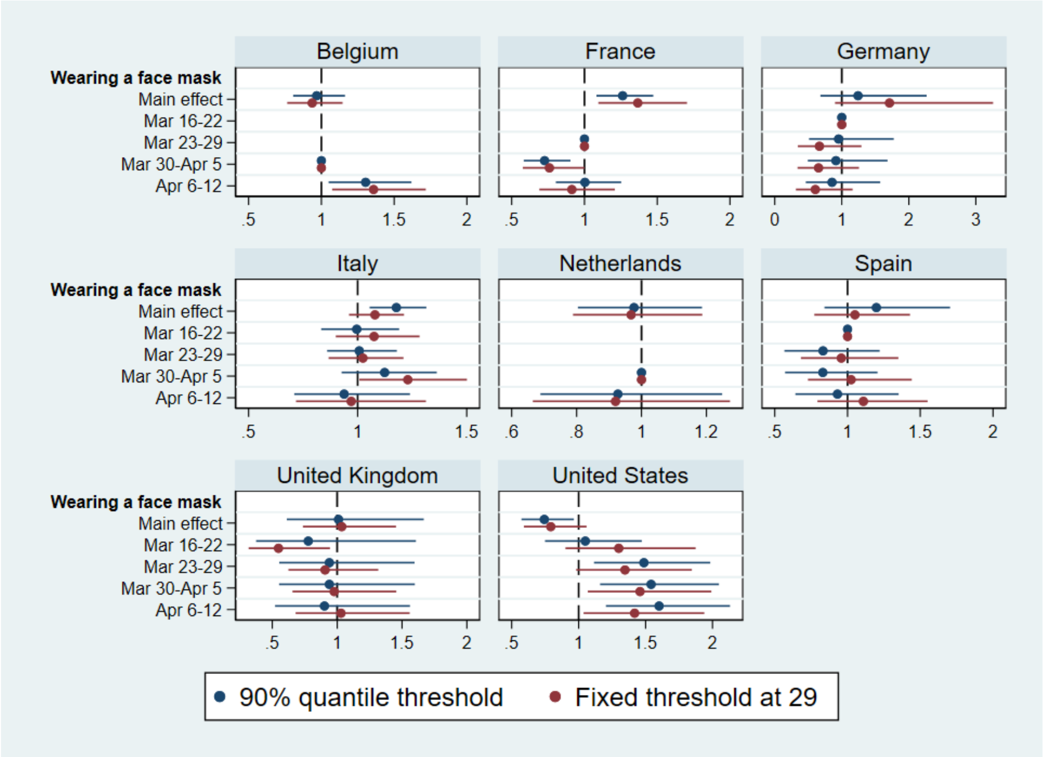


**Figure S14.** Effect of wearing a face mask on overall contacts by study week. Exponentiated coefficients for the protective action "Wearing a face mask", measuring the association between the action and the overall number of social contacts across study weeks. Results for both contact thresholds are shown.

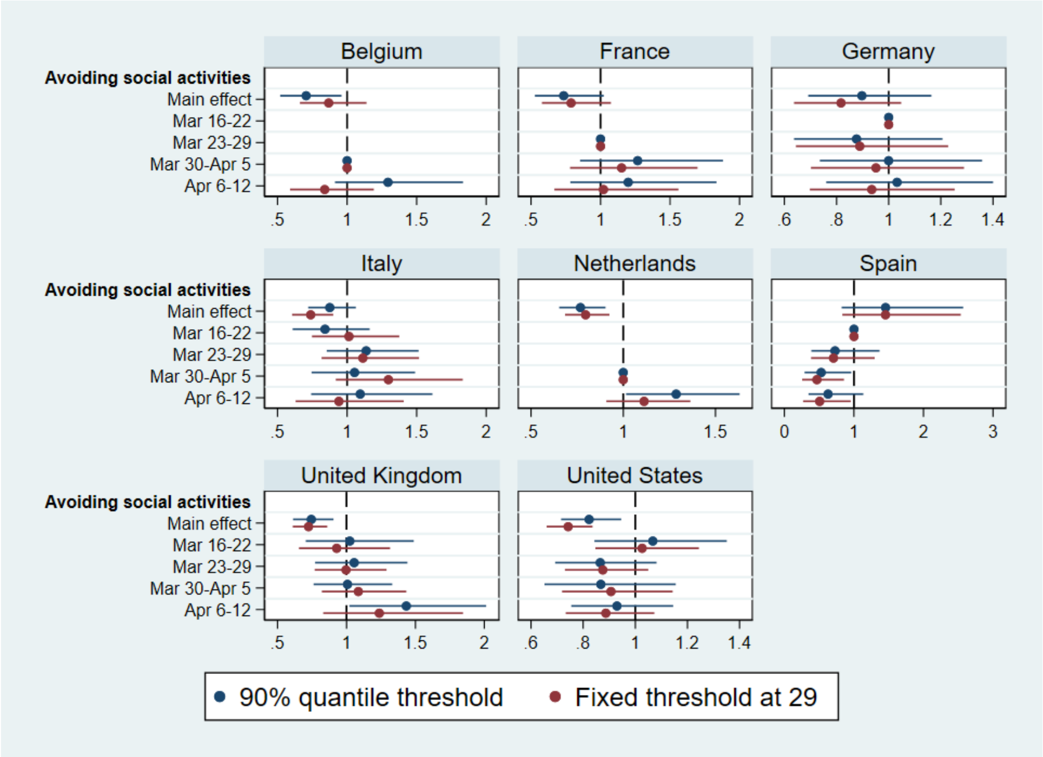


**Figure S15.** Effect of avoiding social activities on overall contacts by study week. Exponentiated coefficients for the protective action "Avoiding social activities", measuring the association between the action and the overall number of social contacts across study weeks. Results for both contact thresholds are shown.

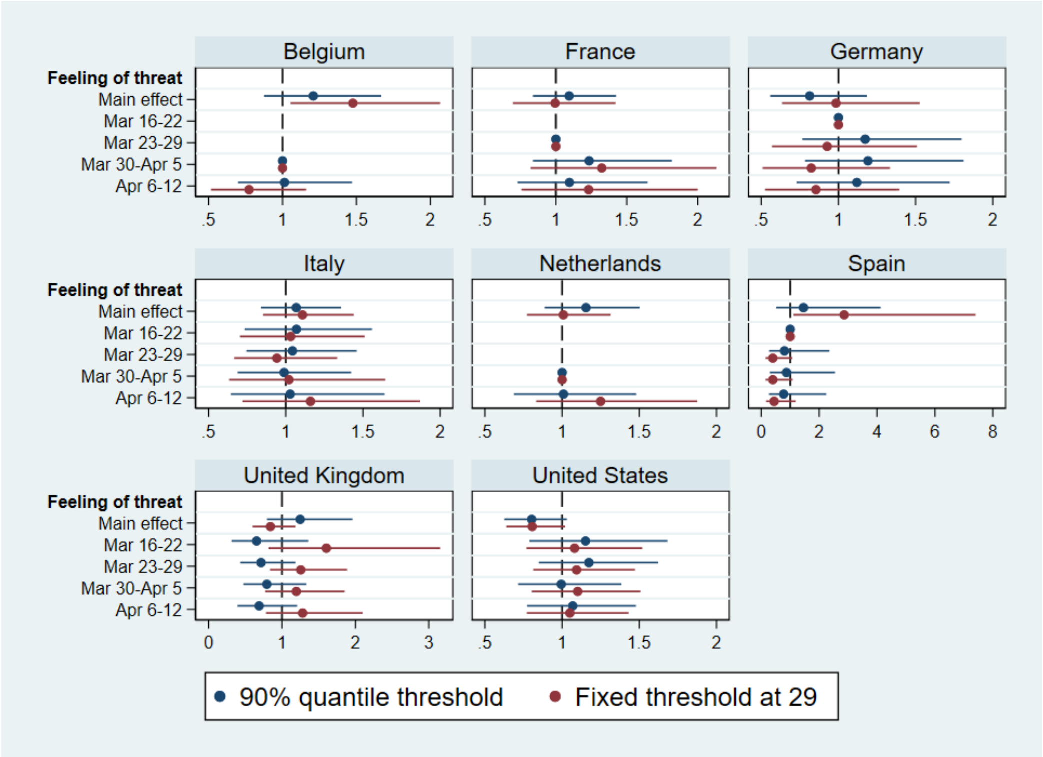


**Figure S16.** Effect of the feeling of threat to oneself and to family on overall contacts by study week. Exponentiated coefficients for the feeling of threat posed by COVID-19 to oneself and the family, measuring the association between the action and the overall number of social contacts across study weeks. Results for both contact thresholds are shown.

### References


1. Perrotta D, Grow A, Rampazzo F, Cimentada J, Del Fava E, Gil-Clavel S, et al. Behaviors and attitudes in response to the COVID-19 pandemic: Insights from a cross-national Facebook survey. medRxiv. 2020;:2020.05.09.20096388.

2. Eurostat, Statistical Office of the European Union. Eurostat regional yearbook: 2019 edition. 2019. http://publications.europa.eu/publication/manifestation_identifier/PUB_KSHA19001ENN. Accessed 15 May 2020.

3. Ipsos. Public Opinion on the Coronavirus outbreak: a multi-country poll from Ipsos. 2020. https://www.ipsos.com/en/coronavirus-public-opinion. Accessed 15 May 2020.

4. Pötzschke S, Braun M. Migrant Sampling Using Facebook Advertisements: A Case Study of Polish Migrants in Four European Countries. Soc Sci Comput Rev. 2016;35:633–53.

5. Hoffman Pham K, Rampazzo F, Rosenzweig LR. Online Surveys and Digital Demography in the Developing World: Facebook Users in Kenya. ArXiv191003448 Cs. 2019. http://arxiv.org/abs/1910.03448. Accessed 1 Dec 2019.

6. Zhang B, Mildenberger M, Howe PD, Marlon J, Rosenthal SA, Leiserowitz A. Quota sampling using Facebook advertisements. Polit Sci Res Methods. 2018;:1–7.

7. Keiding N, Louis TA. Perils and potentials of self-selected entry to epidemiological studies and surveys. J R Stat Soc Ser A Stat Soc. 2016;179:319–76.

8. Mercer AW, Kreuter F, Keeter S, Stuart EA. Theory and Practice in Nonprobability Surveys Parallels between Causal Inference and Survey Inference. Public Opin Q. 2017;81:250–71.

9. US Census Bureau. 2018 Population Estimates by Age, Sex, Race and Hispanic Origin. The United States Census Bureau. https://www.census.gov/newsroom/press-kits/2019/detailed-estimates.html. Accessed 15 May 2020.

10. Mossong J, Hens N, Jit M, Beutels P, Auranen K, Mikolajczyk R, et al. Social Contacts and Mixing Patterns Relevant to the Spread of Infectious Diseases. PLoS Med. 2008;5:e74.

11. Béraud G, Kazmercziak S, Beutels P, Levy-Bruhl D, Lenne X, Mielcarek N, et al. The French Connection: The First Large Population-Based Contact Survey in France Relevant for the Spread of Infectious Diseases. PLoS ONE. 2015;10:e0133203.

12. Teslya A, Pham TM, Godijk NG, Kretzschmar ME, Bootsma MCJ, Rozhnova G. Impact of self-imposed prevention measures and short-term government-imposed social distancing on mitigating and delaying a COVID-19 epidemic: A modelling study. PLoS Med. 2020;17:e1003166.

13. Chu DK, Akl EA, Duda S, Solo K, Yaacoub S, Schünemann HJ, et al. Physical distancing, face masks, and eye protection to prevent person-to-person transmission of SARS-CoV-2 and COVID-19: a systematic review and meta-analysis. The Lancet. 2020;395:1973–87.

14. Diekmann O, Heesterbeek JAP, Metz JAJ. On the definition and the computation of the basic reproduction ratio R0 in models for infectious diseases in heterogeneous populations. J Math Biol. 1990;28:365–82.

15. Wallinga J, Teunis P, Kretzschmar M. Using Data on Social Contacts to Estimate Age-specific Transmission Parameters for Respiratory-spread Infectious Agents. Am J Epidemiol. 2006;164:936–44.

16. Coletti P, Wambua J, Gimma A, Willem L, Vercruysse S, Vanhoutte B, et al. CoMix: comparing mixing patterns in the Belgian population during and after lockdown. Sci Rep. 2020;10:21885.

17. Feehan D, Mahmud A. Quantifying population contact patterns in the United States during the COVID-19 pandemic. medRxiv. 2020;:2020.04.13.20064014.

18. Jarvis CI, Van Zandvoort K, Gimma A, Prem K, CMMID COVID-19 working group, Klepac P, et al. Quantifying the impact of physical distance measures on the transmission of COVID-19 in the UK. BMC Med. 2020;18:124.

19. Klepac P, Kucharski AJ, Conlan AJ, Kissler S, Tang M, Fry H, et al. Contacts in context: large-scale setting-specific social mixing matrices from the BBC Pandemic project. medRxiv. 2020;:2020.02.16.20023754.

20. Arregui S, Aleta A, Sanz J, Moreno Y. Projecting social contact matrices to different demographic structures. PLoS Comput Biol. 2018;14:e1006638.

21. WHO. Transmission of SARS-CoV-2: implications for infection prevention precautions. 2020. https://www.who.int/news-room/commentaries/detail/transmission-of-sars-cov-2-implications-for-infection-prevention-precautions. Accessed 24 Aug 2020.

22. Quinn K. Social media and social wellbeing in later life. Ageing Soc. 2019;:1–22.
